# Lymphotropic Virotherapy Engages DC and High Endothelial Venule Inflammation to Mediate Cancer *In Situ* Vaccination

**DOI:** 10.1101/2025.04.23.25326279

**Authors:** Andrea L. Ludwig, Zachary P. McKay, Griffin P. Carter, Mark A. Katz, Georgia Howell, Vaibhav Jain, Stephanie Arvai, Dirk P. Dittmer, Darell D. Bigner, David M. Ashley, Madison L. Shoaf, Annick Desjardins, Simon G. Gregory, Michael C. Brown, Matthias Gromeier

## Abstract

Intratumor (IT) inoculation of the rhino:poliovirus chimera, PVSRIPO, yielded objective radiographic responses with long-term survival in 20% of patients with recurrent glioblastoma (rGBM). PVSRIPO infects dendritic cells (DCs) and sets up non-cytopathogenic viral (v)RNA replication, which triggers sustained type-I IFN (IFN-I) signaling and antitumor T cell priming. Here we identify IFN-I signaling in glioma-draining cervical lymph nodes (cLN) as a mediator of polio virotherapy. Transient IFN-I signaling after IT therapy was rescued by cervical perilymphatic injection (CPLI) of PVSRIPO, targeting cLN directly. Dual-site (IT+CPLI) PVSRIPO induced profound inflammatory reprogramming of cLN, enhanced vRNA replication and IFN-I signaling in DCs and High Endothelial Venules (HEV), augmented anti-glioma efficacy in mice, and was associated with T cell activation in rGBM patients. A Ph2 clinical trial of IT+CPLI PVSRIPO is ongoing (NCT06177964). This work implicates the lymphatic system as a novel virotherapy target and demonstrates the CPLI concept to complement brain tumor immunotherapy.

## Introduction

Tumor immune surveillance relies on tumor antigen cross presentation and T cell priming by conventional (c)DCs in tdLN,^1–5^ which occur in an IFN-I-dependent manner.^6^ Mere exposure to inflammatory mediators is suboptimal for cDC activation relative to endogenous pattern recognition receptor activation.^7–9^ The impact of IFN-I signaling is notoriously context-dependent:^10^ a signature of sustained, endogenous IFN-I release upon activating [the double-stranded (ds) viral (v)RNA sensor] MDA5 in cDCs/macrophages provided optimal spatiotemporal context for CD8^+^T cell priming.^11,12^ In contrast, IFN-I release upon toll-like receptor agonism in plasmacytoid (p)DCs—mediated by constitutive IFN regulatory factor (IRF) 7 expression in pDCs^13^—contributed little to CD8^+^T cell responses.^12^

Poliovirus pathogenesis studies after oral infection of chimpanzees revealed the lymphatic system as the main viral replication reservoir; virus was found in lymphatic structures at the gastrointestinal (GI) portal of entry (tonsils, Peyer’s patches, mesenteric LN) and in distant axillary or cLN.^14,15^ This corroborated early (from a 1916 polio outbreak) autopsy findings of widespread, profound lymphatic system involvement.^16^ In a systematic study of the (pre-paralytic) phase of poliovirus infection in primates, GI tract CD11c^+^ cells stained positive for viral antigen,^17^ suggesting that poliovirus reaches its lymphatic reservoir via locally infected migratory antigen presenting cells (APCs).

PVSRIPO is the poliovirus type 1 live attenuated (Sabin) vaccine replicating under control of a human rhinovirus type 2 (HRV2) internal ribosomal entry site (IRES), a substitution that mediates profound attenuation^18^ with loss of cytopathogenicity.^19^ Delayed HRV2 IRES-initiated translation of incoming vRNA prevents viral cleavage of the central translation initiation scaffold eukaryotic initiation factor (eIF) 4G and host translation shut-off.^20^ Yet, similar to its progenitors, PVSRIPO succeeds at hijacking host phosphatidylinositol 4-kinase (PI4K) for generating PI4P-enriched vRNA replication organelles.^19,21^ In PVSRIPO-infected APCs, extended vRNA replication generates dsRNA sensed by MDA5^19,22,23^ to induce polar TBK1-IRF3 signaling, and thereby IFN-I release and APC maturation that revives antitumor T cell immunity.^22,24,25^

In this study, we investigated the role of tumor-draining (td)LN in polio virotherapy. We demonstrate that IT PVSRIPO provokes DC homing to tdLN in tumor-bearing mice; induces vRNA replication in tdLN; and that the ensuing tdLN-localized IFN-I response mediates antitumor effects of polio virotherapy. Transcriptomic analyses of cLN in murine glioma revealed that CPLI of PVSRIPO—to target cLN directly—reinvigorates ebbing IFN-I signaling induced by IT PVSRIPO. Successive “dual-site” (IT, then CPLI) targeting with PVSRIPO elevated vRNA replication in cLN far beyond CPLI alone; robustly enhanced IFN-I signaling; mediated profound cLN proinflammatory reprogramming fundamentally distinct from single-site treatment by either IT or CPLI; and improved PVSRIPO efficacy in murine glioma. Spatial analyses of replicating vRNA after dual-site PVSRIPO revealed widespread viral targeting of cLN-resident DCs and endothelial cells in HEV, structures intricately involved in control of adaptive (antitumor) immunity. Based on these studies, three patients with rGBM were treated on Expanded Access protocols with dual-site IT+CPLI PVSRIPO, wherein evidence of post-treatment intratumor T cell inflammation and peripheral expansion of effector CD8^+^ T cells was observed. A Ph2 clinical trial testing this regimen in rGBM (NCT06177964) is accruing patients.

## Results

### PVSRIPO homes to tdLN after intratumor (IT) therapy

We examined if IT PVSRIPO infusion in the brain leads to virus accumulation in tumor/brain-draining cLN—analogous to wild type (wt) poliovirus homing to the lymphatic system after oral infection (**Fig. 1**).^14,15^ To test this, we used mice transgenic for the human poliovirus receptor gene (*PVR*; a.k.a. CD155) (hCD155-tg)^26^ since rodent CD155 homologs do not mediate poliovirus entry. CD155 distribution in hCD155-tg mice and humans is similar;^26^ PVSRIPO-infected hCD155-tg mouse bone marrow-derived DCs (bmDCs) and human monocyte-derived DCs (mdDCs) exhibit similar susceptibility and innate response signatures.^25^ Virus was injected by stereotactic IT inoculation in the syngeneic CT2A mouse glioma model (hCD155-transduced; CT2A^hCD155^) as reported in detail previously (**Fig. 1a**).^27^ Ipsi- (containing tumor/PVSRIPO injection sites) and contralateral brain hemispheres, ipsilateral cLN, ipsilateral kidney, spleen, and serum were analyzed by plaque assay at defined intervals (**Fig. 1b**). Here, and in all subsequent studies, superficial and deep cLN (from individual animals) were pooled to maximize capture of lymphatic drainage.^28^ Kidney is a sentinel tissue, as it “traps” poliovirus passively disseminating with blood, but is not a replication reservoir in chimpanzees^14,15^ or hCD155-tg mice.^29^

**Fig. 1.**
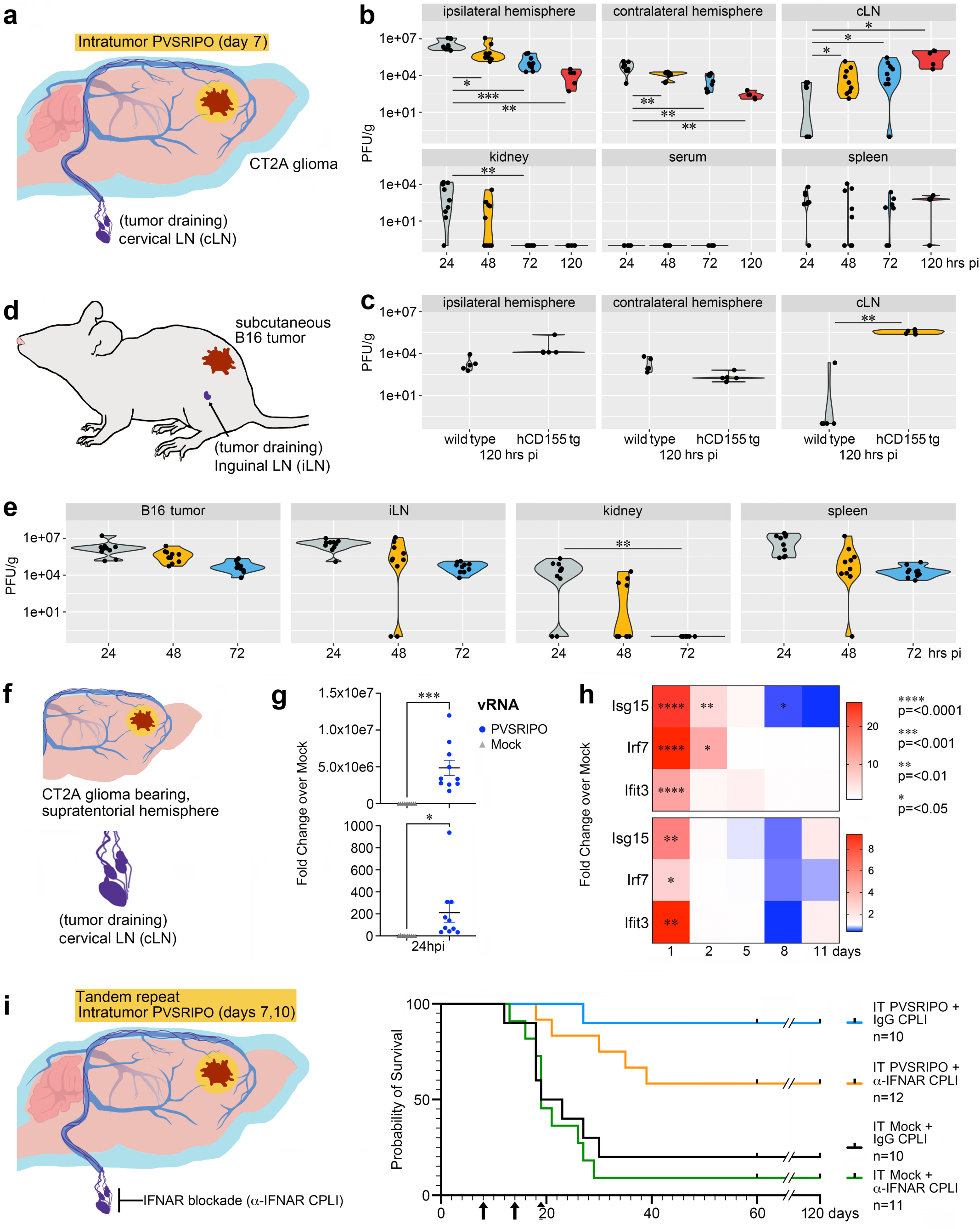
PVSRIPO homes to secondary lymphatic organs after IT treatment in murine CT2A glioma and B16 melanoma. **a**, The murine system of meningeal lymphatic vessels draining to cLN. **b**, Viral titers in the indicated tissue homogenates/serum at days 1-3, 5 post IT inoculation in CT2A glioma [single dose; 5 x 10^7^ plaque forming units (pfu; 5μL)]. The assay was carried out in three repeats with different sampling intervals; data shown are the combined results from two distinct series. **c**, PVSRIPO accumulation in cLN depends on hCD155^+^ host cells. The assay in b was repeated in wt type mice bearing wt CT2A tumors vs. hCD155-tg mice bearing hCD155-tg CT2A tumors. Virus recovery from cLN at day 5 post IT PVSRIPO is shown. **d**, iLN location respective to (subcutaneous) B16 melanoma in mice. **e**, Viral titers in the indicated tissue homogenates at days 1-3 post IT inoculation in B16 melanoma [single dose; 5 x 10^8^ pfu (50μL)]. **f-h**, Tandem repeat IT PVSRIPO (5 x 10^7^ pfu each) vs. mock (vehicle) was administered in CT2A glioma to assess innate host responses in the tumor bearing hemisphere (top row; **f-h**) and ipsilateral cLN (bottom row; **f-h**). RT-qPCR analysis of (+)strand vRNA after the 2^nd^ IT PVSRIPO infusion (**g**). RT-qPCR analysis of IFIT3, ISG15, and IRF7 transcripts 1-11 days after the 2^nd^ IT PVSRIPO infusion (h). The assay was carried out in three independent series with variable test intervals; data shown are aggregate results from all series. **i**, Survival of mice bearing CT2A tumors treated as in **f-h** is diminished by CPLI of anti-IFNAR antibody (400μg) on days 8, 14, and 19 (arrows) vs. CPLI of isotype-matched IgG control.

Virus recovery from the ipsilateral, tumor-bearing hemisphere significantly declined by day 5 (∼1,000-fold vs. day 1), consistent with earlier reports,^22,30^ indicating absent or minimal viral replication in inoculated hCD155^+^ tumors. Abundant virus recovery from contralateral (non-injected) brain declined in step with the opposite hemisphere (**Fig. 1b**). In contrast, we observed significant virus accumulation in (ipsilateral) cLN, where virus titers increased moderately, but steadily (**Fig. 1b**; day 5 p=0.016). This was not due to passive spread with blood, as low-level virus trapping in kidney disappeared by 72h, and because there was no viremia (**Fig. 1b**). Virus isolation from spleen until day 5 may indicate virus homing analogous to wt poliovirus distribution in chimpanzees;^15^ however—in contrast to cLN—splenic viral titers were very low and inconsistent (**Fig. 1b**). To assess viral cLN homing mechanisms, we tested virus distribution in wt mice bearing CT2A^wt^ tumors (non-permissive to PVSRIPO) vs. hCD155-tg mice bearing CT2A^hCD155^ tumors (**Fig. 1c**). Similar virus abundance in the brains of wt vs. hCD155-tg mice emphasized absent viral replication in the tumor. However, virus levels in cLN of wt vs. hCD155-tg mice differed significantly (**Fig. 1c**; p=0.0097), indicating that virus cLN homing is hCD155/host cell-dependent. Biodistribution tests in B16 flank melanomas (hCD155-tg mice; **Fig. 1d**) confirmed abundant virus recovery from iLNs/spleen (relative to sentinel kidney; **Fig. 1e**). In aggregate, our investigations showed that IT administered PVSRIPO homes to and accumulates in tdLN.

### IT PVSRIPO induces LN-localized IFN-I responses that contribute to anti-glioma efficacy

Non-cytopathogenic PVSRIPO infection triggers sustained IFN-I release in primary myeloid cells^19,22,24,25^ and in myeloid infiltrates of GBM *ex vivo* slices.^22^ Thus, we examined IFN-I responses in tumor-bearing hemispheres and ipsilateral cLN after IT PVSRIPO in CT2A glioma (**Fig. 1f-h**). At 24h post IT PVSRIPO, there was significant presence of vRNA in the brain (p=0.0002) and cLN (p=0.027; **Fig. 1g**), accompanied by significant IFN-stimulated gene (ISG) induction (**Fig. 1h**). This was accompanied by increases in LN weight, GzmB^+^ CD8^+^ T cells, CD11c^+^ MHC II^+^ DCs, and a significant decline in Tregs in cLN (Supplementary Fig. 1a; p=0.013). The IFN-I signature dissipated by ∼5 days in the brain and much earlier, ∼2 days, in cLN (**Fig. 1h**). In fact, ISG mRNA levels decreased below the mock controls by ∼day 8 in cLN (**Fig 1h**), likely due to rebound negative feedback responses.^31^ To assess the relevance of cLN IFN-I for polio virotherapy, we tested survival after tandem repeat IT PVSRIPO combined with mock (non-specific IgG) or anti-IFNα/β receptor (IFNAR) antibody delivered by CPLI (**Fig. 1i**). IFNAR blockade diminished the antitumor efficacy of PVSRIPO (**Fig. 1i**). Thus, IFN-I responses in cLN— while transient—contribute to the antitumor efficacy of polio virotherapy.

### PVSRIPO vRNA replicates in tdLN-associated CD11c^+^ cells after IT therapy and induces DC migration from tumor to tdLN

MDA5-TBK1-IRF3 innate signaling drives sustained IFN-I release upon PVSRIPO infection^19,20,22,24,25^ that induces the immunoproteasome, transporter associated with antigen processing (TAP1),^24^ and the MHC I peptide loading complex; CD80/86, CD40 maturation markers;^25^ and C-C chemokine receptor 7 (CCR7),^25^ the LN homing receptor on DCs.^32^ Thus, trafficking of infected DCs/other myeloid cells to tdLN could explain vRNA presence and IFN-I responses after IT PVSRIPO. To test this possibility, we treated CT2A glioma-bearing mice with IT PVSRIPO, harvested cLN single cell suspensions, and either negatively selected DCs or enriched DCs via CD11c^+^ bead-isolation (**Fig. 2a**). RT-qPCR detected (+)strand vRNA in CD11c^+^-bead (positively) selected- and DC (CD3^-^, CD19^-^, NK1.1^-^, F4/80^-^)-negatively selected cell subsets (Supplementary Fig. 1b, c). We confirmed these findings using flow cytometry:hybridization chain reaction-fluorescence *in situ* hybridization (HCR-FISH) to identify vRNA-containing cLN cell subsets after direct cLN targeting via PVSRIPO CPLI in non-tumor bearing hCD155-tg mice. These analyses revealed the presence of vRNA in CD11c^+^ bead isolated- (**Fig. 2a, b**) and in DC-negatively selected (Supplementary Fig. 1c) cLN cell subsets after CPLI of PVSRIPO *in vivo* (**Fig. 2c**). Enrichment of vRNA occurred predominantly in the cLN ipsilateral to the site of CPLI (**Fig. 2c**); detection of (-)strand vRNA implies active PVSRIPO RNA replication in infected cLN cell subsets. This agrees with ISG signatures in cLN after IT PVSRIPO (**Fig. 1h**), and our finding that innate MDA5-TBK1-IRF3 signaling is triggered by ds vRNA.^19,22^

**Figure 2.**
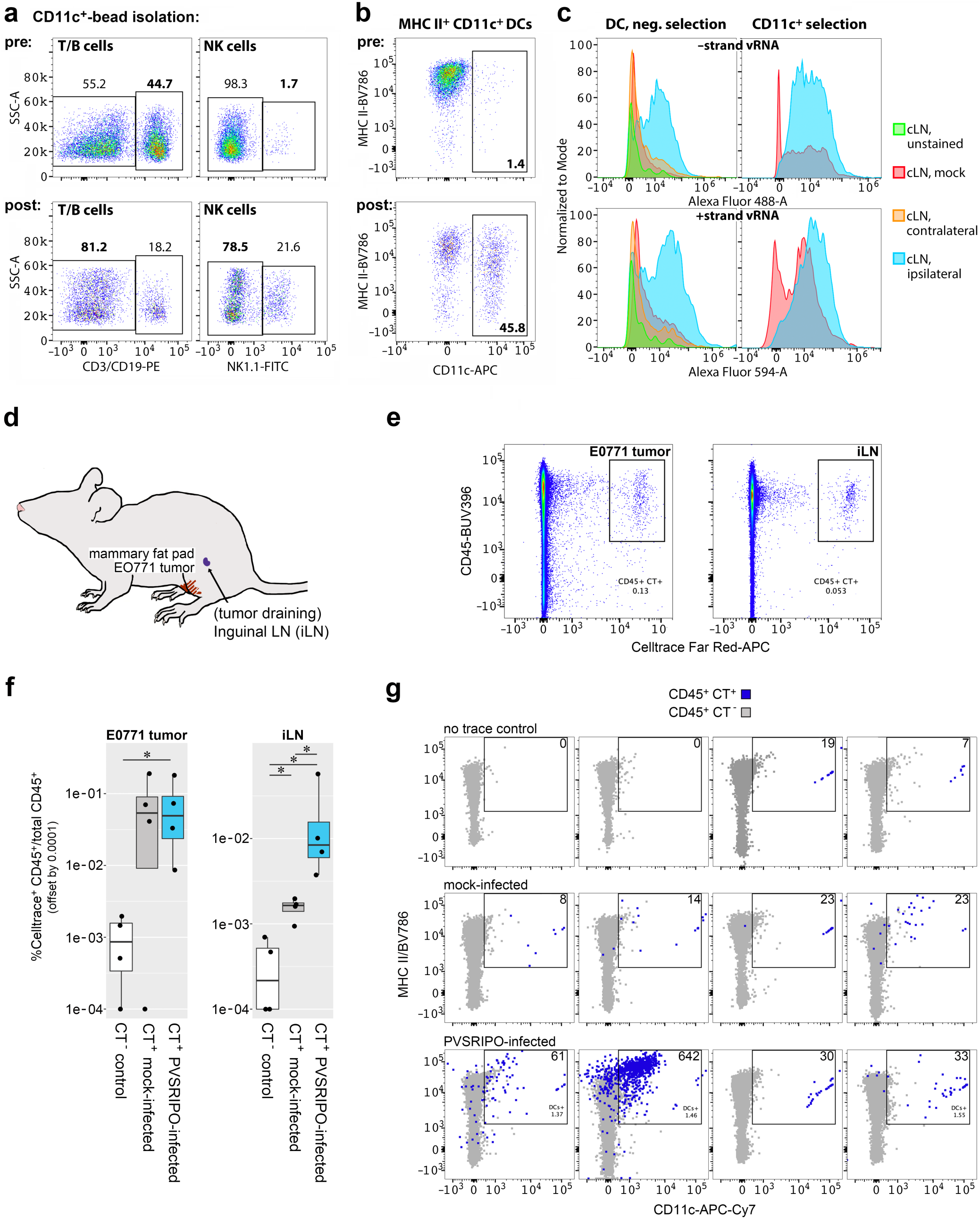
Intratumor PVSRIPO induces transient IFN-I responses in the tumor-bearing hemisphere and cLN. **a**, **b**, Depletion of T/B cells, and enrichment of (CD11c^+^) NK cells (**a**) and MHC II^+^ CD11c^+^ DCs (**b**) after CD11c^+^-bead isolation of cells from dissociated cLN. **c**, Flow cytometry:hybridization chain reaction-fluorescence *in situ* hybridization (HCR-FISH) of (-) and (+)strand vRNA in MHC II^+^ CD11c^+^ cells. Cell subsets from ipsi- or contralateral cLN of mock vs. PVSRIPO-treated mice were isolated by DC-negative selection (see Supplementary Fig. 1b) or by CD11c^+^-bead isolation (**a**, **b**). **d**, DC homing after adoptive transfer in the orthotopic (mammary fat pad) E0771 mouse breast cancer model. **e**, bmDCs were harvested from hCD155-tg mice, differentiated with FLT3 ligand,^25^ and treated with PVSRIPO or vehicle *in vitro* (24h), labeled with Celltrace far red (CT^+^) fluorescent marker immediately after, and adoptively transferred into E0771 tumors. Tumors and iLN were harvested (16h) and analyzed by flow cytometry to detect trace-labeled cells. A pilot feasibility assay to detect (PVSRIPO infected) CT^+^ cells in tumor/iLN suspensions from one mouse is shown. **f**, CT^+^ cells in E0771 tumor (left) and iLN (right) 16h after IT transfer of unlabeled, untreated bmDCs (n=4 mice), or labeled bmDCs treated with PVSRIPO (n=4 mice) vs. mock (n=4 mice). **g,** iLN from all 12 mice in the assay in **f**/**g** were processed into cell suspensions and examined by flow cytometry (see Supplementary Fig. 1d for gating strategy) to filter out CD3^+^/CD19^+^/NK1.1^+^/CD11b^+^/CD14^+^ cells; gating for MHC II^+^ CD11c^+^ cells/CT^+^ cells in this compartment are shown. The assay was repeated in three independent series.

We next tested if PVSRIPO-infected DCs traffic from the tumor to tdLN by adoptive transfer of Cell-Trace-labeled (CT^+^) bmDCs into orthotopic E0771 tumors and tracking CT^+^, mock-vs. PVSRIPO-infected DCs in iLN (**Fig. 2d, e**). We chose orthotopic E0771, rather than CT2A glioma, to overcome volume restrictions with intracranial delivery that limit the number of DCs that could be transferred. While CT^+^ cell recovery from E0771 tumors did not differ between the mock- and PVSRIPO cohorts, there was significantly increased recovery of PVSRIPO-vs. mock-infected CT^+^ cells from iLN (**Fig. 2f**; p=0.028). Gating for MHC-II and CD11c confirmed that CT^+^ cells were indeed DCs (**Fig. 2g**). Thus, PVSRIPO infection enhances DC migration and LN homing from the tumor, possibly explaining the induction of IFN-I responses within tdLN after IT PVSRIPO.

### Combined, successive (dual-site) PVSRIPO IT and CPLI mediate profound inflammatory reprogramming of cLN

Both deep and superficial cLN are sites of brain-derived epitope presentation,^33,34^ and are implicated in cDC1-dependent^35^ anti-glioma T cell priming.^36^ We discovered that IT PVSRIPO induces cDC tdLN homing, vRNA replication and IFN-I signaling in cLN that mediate antitumor efficacy. Yet, IFN-I responses are transient and restrained by negative feedback. Since drug delivery to cLN via CPLI is clinically feasible,^37,38^ we hypothesized that PVSRIPO CPLI may sustain and boost beneficial IFN-I responses in cLN after IT therapy. For unbiased tests of this hypothesis, we compared cLN transcriptomes from four cohorts of CT2A glioma-bearing mice treated with mock or IT PVSRIPO, CPLI, or both as depicted (**Fig 3a, b**).

**Figure 3.**
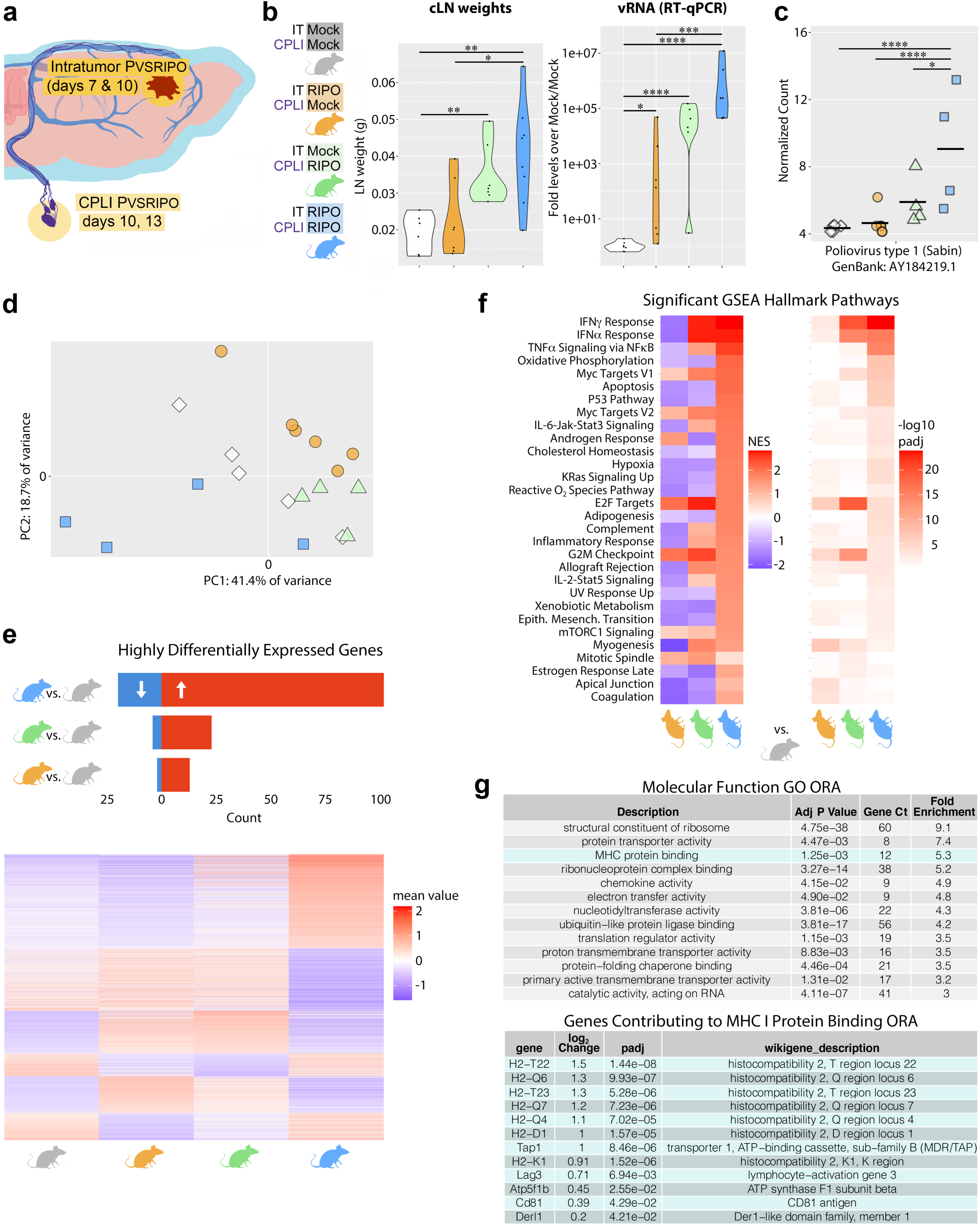
Synergistic effect of PVSRIPO dual-site treatment in tumor and cLN. **a**, **b**, Mice were treated at defined intervals post tumor implantation with PVSRIPO (**a**) in 4 groups where IT (tandem repeat infusion with 5 x 10^7^ pfu) + CPLI (10^7^ pfu at each interval) of PVSRIPO vs. mock was combined variably as shown (**b**). cLN were harvested from mice in each of the four cohorts, weighed (left), and processed for RNA isolation and RT-qPCR of (+)strand vRNA (right). **c**, The frequency of (+)strand vRNA as detected by RNA-seq. See Figure 1g for a legend of statistical significance. **d**, Principal component analysis (PCA) showing the first two principal components of all samples after removing outliers and correcting for batch effects demonstrate clustering by treatment group. **e**, The number of highly differentially expressed genes (padj < 0.05 and absolute log2 Fold Change value > 2) in cLN of each treatment group (IT, CPLI, dual-site PVSRIPO; see **b** for color code) compared to mock (top). Heatmap of all differentially expressed protein-coding genes in any of the three treatment groups compared to mock organized by high-to-low expression of each gene differentially expressed in dual-site, CPLI, and IT treatment groups (bottom). Volcano plots of these comparisons are shown in Supplementary Figure 2a-c. **f**, GSEA hallmark pathway (padj < 0.05 for at least one comparison) enrichment and adjusted p-value by each treatment group (compared to mock) as organized by normalized enrichment score (NES) for pathways significantly enriched in the dual-site PVSRIPO-, single-site CPLI- or IT cohorts, compared to the dual-site mock group (see Supplementary Figure 2d-f for extended data). **g**, Gene ontology overrepresentation analysis using molecular function terms of significantly upregulated genes in dual-site PVSRIPO-treated mice compared to dual-site mock (top); genes contributing genes to MHC protein binding term being found as overrepresented (bottom). For comparison of single-site IT/CPLI PVSRIPO vs. mock see Supplementary Figure 2g, h.

Dual-site treatment (IT+CPLI) produced the largest increase in cLN weight and vRNA content (per RT-qPCR) amongst the four groups (**Fig. 3b**); cLN vRNA abundance (per RNA seq) was significantly increased with dual-site treatment compared to CPLI alone (**Fig. 3c**; p=0.001), indicating that PVSRIPO-infected myeloid cell homing may “prime” cLN for subsequent virus challenge via CPLI. Transcriptomes largely clustered by group with dual-site treatment cLN appearing distinct from other cohorts when analyzed by the 1^st^ and 2^nd^ principal components (explaining ∼60% of variance within samples; **Fig. 3d**). Differential expression analysis (vs. mock) revealed unique activation patterns across each of the four groups, indicating that dual-site treatment did not merely yield additive gene expression changes, but rather engaged distinct inflammatory programs in cLN (**Fig. 3e**).

Gene set enrichment analysis (GSEA) revealed robust IFNα/γ pathway gene expression enrichment after PVSRIPO CPLI (regardless of IT treatment)—which was suppressed in cLN after IT PVSRIPO alone (**Fig. 3f**)—consistent with ISG RT-qPCR analyses (**Fig. 1h**). This confirms that PVSRIPO CPLI rescues and sustains IFN-I signaling in cLN after IT virus administration. Moreover, dual-site treatment led to enrichment of multiple pathways not observed in either monotherapy (IT or CPLI) group (**Fig. 3f**), eg. TNF signaling and metabolic process alterations. Gene ontogeny (GO) analysis revealed significantly enriched transcripts after dual-site PVSRIPO (relative to dual-site mock) that included MHC protein binding and chemokine activity only in the former (**Fig. 3g**). This is consistent with induction of antigen processing, loading and cross presentation machinery in PVSRIPO-infected APCs.^22,24,25^ Immune phenotyping of ipsilateral cLN after dual-site PVSRIPO revealed decreases in Tregs, and significantly increased MHC II expression in monocytes (vs. all treatment groups) and cDC frequency (vs. mock) (Supplementary Fig. 3a). Together these data indicate that combining IT and CPLI treatment with PVSRIPO may serve to boost IFN-I signaling and antigen presentation in cLN.

### PVSRIPO vRNA replicates in cLN CD11c^+^ cells and high endothelial venules (HEV) after combined IT and CPLI

Our data imply that IT PVSRIPO targets DCs in tdLN (**Fig. 2**) and that CPLI enhances inflammatory reprogramming of cLN after IT treatment (**Fig. 3**). We sought to define the spatial context of vRNA replication and IFN signaling in tdLN after dual-site PVSRIPO (**Fig. 4**). HCR-FISH for (-)strand vRNA marks cells harboring active vRNA replication and avoids detection of input (+)strand genomic RNA in cLN tissue sections (**Fig. 4a**). It revealed CD11c^+^ cells in cLN follicles, presumably DCs, co-staining for (-)strand vRNA (**Fig. 4b**), as suggested by other results in this report (**Fig. 2**) and prior observations of PVSRIPO-infected cells in LN.^25^ These findings reverberate with prior data showing poliovirus antigen in CD11c^+^ APCs in the periphery after oral infection of non-human primates with wt poliovirus.^17^

**Figure 4.**
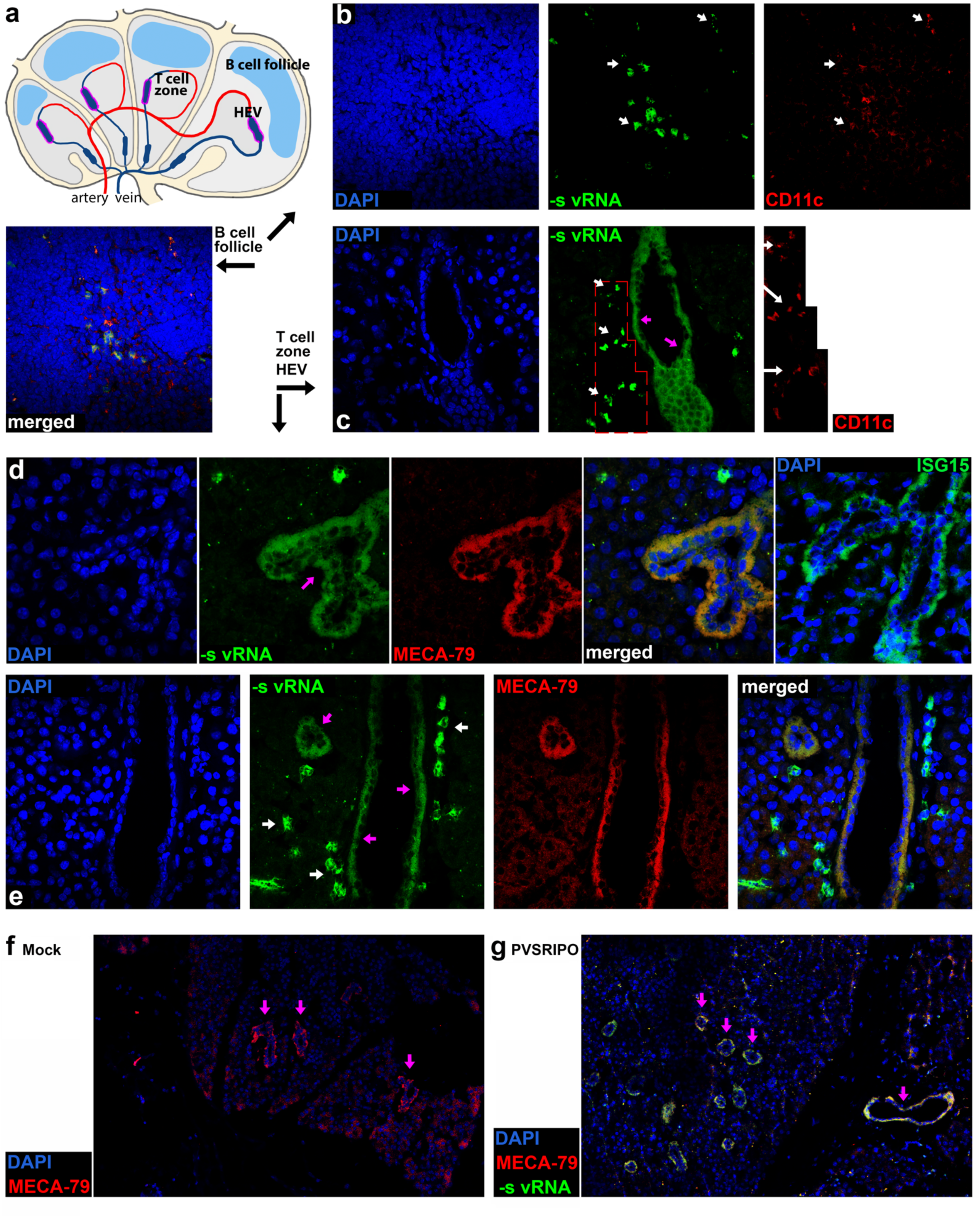
PVSRIPO targets CD11c^+^ DCs and MECA-79^+^ HEV within cLN after dual-site treatment of glioma-bearing mice. **a**-**g**, Mice were treated with dual-site PVSRIPO (see Figure 3a), euthanized 24h after the 2^nd^ CPLI of PVSRIPO (day 14), and ipsilateral cLN were removed for (-)strand vRNA HCR-FISH and immunofluorescence analysis. Anatomical context of LN areas of interest emerging from our studies: B cell follicles (**b**) and HEV (**c**-**g**). **b**, Fresh frozen cLN were cryosectioned (7μm), formaldehyde-fixed and processed for staining as shown. cLN follicles contained distinct groups of cells that stained (-)strand vRNA^+^ and CD11c^+^. **c**, In adjacent areas, (-)strand vRNA^+^ structures reminiscent of HEV, surrounded by (-)strand vRNA^+^ CD11c^+^ cells were observed. The marked area in the HCR-FISH-stained section is shown enlarged and stained for CD11c to the right. **d**, The identity of (-)strand vRNA^+^ structures reminiscent of HEV was determined by co-staining with MECA-79; HEV displayed active IFN-I responses (ISG15). **e**, (-)strand vRNA^+^ MECA-79^+^ HEV surrounded by (-)strand vRNA^+^ cells—likely DCs—lining up at their exterior surface. **f**, **g**, Low magnification images show disseminated, enlarged (-)strand vRNA^+^ MECA-79^+^ HEV, suggesting disseminated infection of HEV.

Intriguingly, (-)strand vRNA was also detected in HEV (**Fig. 4c**), critical gatekeepers of adaptive immunity by controlling T cell traffic into and out of LNs.^39^ HEV identity was confirmed by co-staining with the HEV-specific marker, MECA-79 (**Fig. 4d**).^40^ Beyond (-)strand vRNA, MECA-79^+^ cells stained positive for ISG15, identifying HEV as sites harboring active antiviral IFN-I signaling (**Fig. 4d**). DCs homing to LN were proposed to control HEV endothelial cell proliferation, thus regulating HEV abundance and size, and T cell influx, dwell time and egress.^41^ We observed (-)strand vRNA^+^ CD11c^+^ cells congregating around and lining up in parallel to (-)strand vRNA^+^ HEV (**Fig. 4c, e**). MECA-79^+^ (-)strand vRNA^+^ staining was evident in most, if not all, HEV throughout the cLN, and HEV number and size were increased compared to cLN of mice in the dual-site mock cohort (**Fig. 4f, g**). MECA-79^+^ HEV cells co-stained for (transgenic) hCD155, indicating that they are plausible viral targets (Supplementary Fig. 4). Disseminated vRNA replication in cLN occurs despite PVSRIPO’s non-cytopathogenic phenotype that precludes lytic spread of virus. A possible explanation is non-lytic viral spread of poliovirus via phosphatidylserine lipid-enriched vesicles,^42^ which also occurs with PVSRIPO (Supplementary Fig. 5). Thus, dual-site PVSRIPO therapy produces sustained IFN-I responses, boosts antigen presentation gene expression, and engages CD11c^+^ DC and HEV inflammation in cLN.

### CPLI enhances antitumor activity of IT PVSRIPO, is feasible in rGBM patients, and engages T cell activation and effector function in humans

Dual-site treatment with PVSRIPO induces superior cLN proinflammatory engagement. Accordingly, combined IT and CPLI of PVSRIPO generated augmented antitumor effects in mice bearing CT2A gliomas, with 4/10 surviving >60 days in the combination group vs 1/10 after IT therapy alone (**Fig. 5a**). CPLI alone failed to extend survival in mice, indicating that IT therapy must be implemented to engage the local glioma microenvironment. Similarly, in subcutaneous B16 melanoma, we observed that intranodal (IN) therapy—where Evans blue tracing was used to directly target the inguinal LN (tdLN in this model; **Fig. 1d**)^43^— alone mediated an early antitumor effect but was less effective than IT administration at delaying tumor growth (**Fig. 5b**). Together, these data support targeting tdLN to augment IT polio virotherapy.

**Figure 5.**
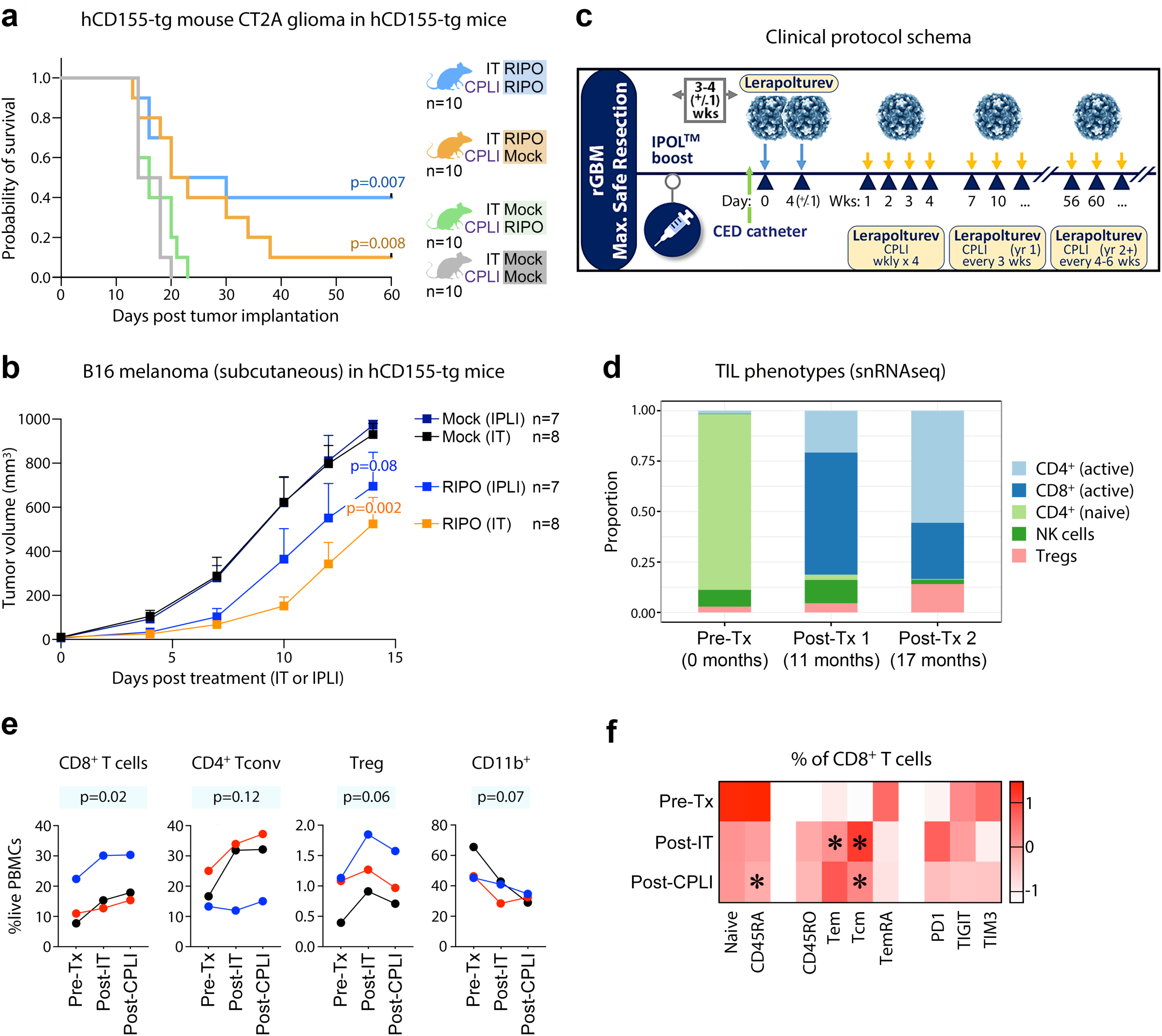
CPLI enhances antitumor activity of IT PVSRIPO, is feasible in rGBM patients, and engages T cell activation and effector function in humans. **a**, hCD155tg mice bearing intracranial CT2A^hCD155^ gliomas were treated IT with mock or PVSRIPO (2.5 x 10^7^ pfu) 7 and 10 days after tumor implantation and either mock or PVSRIPO (10^7^ pfu) CPLI on days 10, 13, and 16. Survival is shown, p-values are from Mantel-Cox log-rank test. **b**, hCD155-tg mice bearing subcutaneous B16^hCD155^ tumors were treated with mock or PVSRIPO (3 x 10^8^ pfu) IT or inguinal perilymphatic injection (IPLI) 8 and 12 days after tumor implantation. Tumor volume -/+ SEM is shown; p-values are from two-way ANOVA vs respective (IT or IPLI) mock control. **c**-**f**, Patients with histologically confirmed rGBM (n=3) received tandem repeat IT and iterative CPLI clinical-grade PVSRIPO (Lerapolturev) according to the schema shown (**c**). **d**, Predicted T/NK cell subset proportions from longitudinal single-nuclei RNAseq of tumor tissue from one patient pre-treatment (Pre-Tx), 11-months post tandem repeat treatment (Post-Tx 1), and 6 months after a second tandem repeat IT treatment (Post-Tx 2; 17 months after first IT infusion). See Supplementary Table 2 and Supplementary Fig. 6a-c for extended data. **e**, Peripheral blood cell proportions determined by flow cytometry (see Supplementary Fig. 6d, e for representative gating) at Pre-Tx (time of surgery, **c**), Post-IT (4 days after the 1^st^ IT infusion), and Post-CPLI (8 weeks after the 1^st^ IT infusion); each line/color represents a different patient; p-values from one-way ANOVA for each feature. **f**, Mean percentage of CD8^+^ T cells with indicated phenotypes in peripheral blood at the same time points as **e**; (*) denotes Bonferroni adjusted paired t-test p<0.025 versus pre-tx time point.

We developed a randomized Ph2 clinical protocol in which survival of patients with rGBM receiving dual-site [tandem repeat IT PVSRIPO followed by iterative CPLI of PVSRIPO (clinical drug name Lerapolturev); (**Fig. 5c**)] will be compared to that of patients treated with lomustine. Prior to initiation of the now ongoing Ph2 trial (NCT06177964), three patients with rGBM were treated per the protocol under separate Expanded Access INDs and IRB/FDA-approved protocols. The combined infusions were well tolerated in all three patients. Glioma tissue from one patient pre- and post dual-site IT and CPLI PVSRIPO was analyzed by single nuclei RNAseq, revealing increased activated CD4^+^ and CD8^+^ T cell signatures (**Fig. 5d**). Moreover, increased proportions of peripheral CD8^+^ T cells were observed after completion of IT therapy (day 5) that further increased after CPLI injections in all 3 patients (6 weeks after initiating CPLI; **Fig. 5e**). Analysis of immune cell phenotypes revealed significant increases in T effector and central memory CD8^+^ T cell phenotypes, without increases in exhaustion markers TIGIT and TIM3 (**Fig. 5f**). These data corroborate activation of CD8^+^ T cells within the tumor site and imply that combined IT and CPLI delivery of PVSRIPO engages CD8^+^ T cell activation.

## Discussion

Cytotoxic CD8^+^ T cell immunity is quintessentially an antiviral response against categorically intracellular pathogens; it depends on APCs consolidating a range of orchestrated contextual signals^44^ that only natural virus infection may provide.^7^ Yet, applying a “virotherapy-as-APC-stimulant” concept is hindered by the fact that virus:host co-evolution selected for variants with elaborate host immune interference ploys that overwhelmingly target APCs. Suppression of IFN-I, peptide transport/MHC I loading, MHC I expression/trafficking, LN migration/homing, antigen cross presentation, maturation, T cell priming and outright viability are common viral host immune interference tactics.^45–50^ Poliovirus is no exception. It uses extremely harsh measures of host interference via immediate eIF4G cleavage with host protein synthesis shut-off ^51^ and host defense blockade. The investigations reported here imply that wild type poliovirus may specifically target migratory cDCs and HEV in LN—a central nexus of the adaptive immune response—for this destructive process. By generating PVSRIPO—ablating the virus’ ability to execute eIF4G cleavage^19,20^—we created a form of poliovirus that retains target specificity for migratory cDCs, tdLN, LN cDCs and HEV, but neither prevents nor counters host immunity.

PVSRIPO triggers polar MDA5-TBK1-IRF3 innate signaling in APCs; HCR-FISH detection of (-)strand RNA in DCs/HEV implies active vRNA replication/the presence of dsRNA. It extends observations in *in vitro* systems^19^—that despite deficits in subduing host cells, PVSRIPO is capable of establishing vRNA replication—to its natural reservoir *in vivo*. Consistent with a non-cytopathogenic phenotype,^19,20,22,24,25^ and lacking pathogenicity in IND-directed primate toxicology,^52^ PVSRIPO did not cause cytopathogenic changes in cLN, to HEV or to DCs; rather our evidence suggests HEV expansion, both in numbers and size. Since revealing the lymphatic system as the main replication reservoir in the 1950s,^14,15^ the identity of poliovirus target host cells in this organ remained elusive. We only recently identified cDCs and HEV endothelial cells as sites for PVSRIPO RNA replication and dsRNA-triggered sustained IFN-I release in LN; much research effort beyond the scope of this study is required to decipher the likely complex immunological implications of this target specificity.

Single intratumor infusion of PVSRIPO in the necrotic core of rGBM has generated promising clinical results with durable radiographic responses and long-term survival in a subset of ∼20% of patients.^53^ Other recombinant viral agents have been tested in this indication with similar delivery regimens and also yielded intriguing responses in select patients.^54–56^ Intuitively, a single infusion into the necrotic, hypoxic core of the deeply immune-suppressive and -skewing rGBM microenvironment may be sub-optimal for generating clinically effective tumor immune surveillance in the brain. Promising steps to improve virus delivery methods, e.g. iterative IT administration, have been proven safe.^57^ Yet, the unique immune-stimulatory potential of the antiviral immune response, while well supported by preclinical work, has yet to demonstrate compelling clinical promise. Here we discovered means to engage the lymphatic system in the brain with a naturally lymphotropic, highly neuroinvasive agent through viral targeting and ensuing antiviral immune responses in cLN that support antitumor efficacy of IT virotherapy. CPLI of PVSRIPO alone failed to produce antitumor effects in murine glioma. Rather, our RNAseq studies specifically imply dual-site treatment, potentially integrating PVSRIPO-infected migratory DC traffic from the (IT-infused) tumor and the (CPLI) periphery, in proinflammatory reprogramming of cLN. Iterative CPLI affords the opportunity to deliver multiple repeat immunotherapy without repeated invasive neurosurgical procedures. Tandem repeat IT polio virotherapy with adjuvant iterative CPLI of PVSRIPO was well tolerated in rGBM patients and was associated with evidence of activated CD8 T cells within the tumor and periphery.

Pre-existing anti-polio immunity is near-universal, all patients treated in the Ph1 study of PVSRIPO were confirmed seropositive for neutralizing anti-polio antibodies, and a dose of the standard pediatric polio vaccine in the US (trivalent inactivated Salk vaccine) is administered at least 1 wk prior to IT PVSRIPO infusion in all subjects.^53^ Pre-treatment poliovirus-specific antibody titers were associated with longer survival after PVSRIPO therapy in patients with rGBM, and pre-existing anti-polio immunity augmented PVSRIPO antitumor efficacy in murine melanoma and breast cancer models.^58^ Survival assays in the CT2A murine glioma model reported here were carried out in hCD155-tg mice immunized against poliovirus, as previously reported.^58^ While the contribution of pre-existing anti-polio immunity was not directly tested in this study, we conclude that dual-site PVSRIPO exhibits vRNA replication phenotypes and the antiviral/IFN-I responses they elicit, in the presence of pre-existing anti-poliovirus immunity. Our collective findings support the development of an ongoing, randomized Ph2 clinical trial to test the efficacy of this approach. They also imply that combining CPLI of virotherapy and/or other *in situ* vaccine approaches with intratumor delivery may augment their antitumor T cell engaging capacity.

## Data Availability

All data produced in the present study are contained in the manuscript and transcriptomic raw data will be shared via a public repository.

## Acknowledgements.

We thank N. Altan Bonnet (NIH, NHLBI) who provided valuable technical assistance with EV isolation, anti-poliovirus capsid antibody A12, and expert review of FISH imaging results. We thank R. McNamara (currently Harvard University) for sharing equipment and providing technical advice on EV isolation and analysis procedures. We thank E. Dobrikova, M. Burnett and D. Satterfield (Duke University) for help with experimental procedures.

## Methods

### Ethics Statement

Human correlative analyses were reviewed and approved by the Institutional Review Board (IRB) at Duke University School of Medicine (Pro00114031; Pro00113348; Pro00115266).

### Mice

Wt C57Bl/6 mice were obtained from Jackson Laboratory. Homozygous hCD155-tg mice^26^ were provided generously by S. Koike (Tokyo Metropolitan Institute of Medical Science, Tokyo, Japan) and maintained as a breeding colony at the Duke University Laboratory Animal Resource. Male and female mice were used at 8-12 weeks of age. Mice were housed in the Duke University Cancer Center Isolation Facility under Biosafety Level 2 (BSL2) conditions with 12-hour light/dark cycles, relative humidity of 50 ± 20%, and temperature of 21 ± 3°C.

### Virus, Cells, Antibodies Used *In Vivo*

PVSRIPO was propagated in HeLa R19 cells, purified, and quantified by plaque assay as described earlier.^20^ We used “plain” mouse CT2A glioma (kindly provided by P. Fecci, Duke University School of Medicine, Durham, NC, USA), mouse E0771 breast cancer (kindly provided by G. Palmer, *ibid*.), mouse B16.F10 melanoma cells (ATCC) and the hCD155-tg transduced variants CT2A^hCD155^ and B16^hCD155^ (all in the C57Bl/6 genetic background) for implantation into wt or hCD155-tg C57Bl/6 mice. For hCD155-transduction, cells were infected with a lentiviral hCD155 expression vector and selected through fluorescence-activated cell sorting (FACS) with αCD155-PE (BioLegend).^27^ We used human A375 melanoma cells (kindly provided by D. Tyler, Univ. of NC at Chapel Hill) for isolation of extracellular vesicles (see below). For tumor implantation, cells were propagated in Dulbecco’s Modified Eagle Medium (DMEM) (Invitrogen) with 10% fetal bovine serum (FBS; Sigma, #F0926) to 60-70% confluency for harvest. For *in vivo* IFNAR blockade, 400μg of anti-mouse IFNAR1 (BioXcell; InVivoMAb #BE0241) or isotype-matched control mouse IgG1 with unknown specificity (BioXcell; InVivoMAb #BE0083) were used for CPLI in mice as shown.

### Tumor Models, Adoptive Transfer

All animal procedures were performed under a Duke University School of Medicine IACUC-approved vertebrate animal use protocol. Where indicated, anti-poliovirus immunization occurred 45 and 31 days prior to tumor administration with two IM injections of PVSRIPO diluted in phosphate buffered saline (PBS; Gibco) to 2 x 10^7^ pfu/ml and mixed 1:1 with Alhydrogel (Invitrogen) with one (50μl) injection in each hind quad.^58^ Stereotactic orthotopic implantation of mouse glioma and stereotactic virus/mock inoculations are described in detail elsewhere.^27^ PVSRIPO/mock IT treatment in the brain was performed on day 7 after tumor implantation; tandem repeat treatment was administered on days 7 and 10. Doses used are indicated in figure legends; all stereotactic infusions delivered a volume of 5μl. For survival experiments, individual mice were divided into treatment groups randomly. CPLI of PVSRIPO/ mock was delivered as a 50μl subcutaneous injection on the side of the neck ipsilateral to the tumor implantation site with a 28G Insulin Syringe (BD #329461). For B16 melanoma implantation, B16 cells diluted in PBS (5×10^5^ cells; 50μl) were injected subcutaneously into (previously shaved) flanks of mice. E0771 breast cancers were orthotopically implanted into the 4^th^ mammary fat pad (5×10^5^ cells; 50μl).^58^ PVSRIPO treatment was administered IT at the doses indicated in figure legends. Tumor volumes were measured using Vernier calipers (Thermo-Fisher) every 48h and calculated using the equation L × W × W/2. Mice were euthanized when tumors reached a volume of 1000 mm^3^ or the indicated experimental endpoint. For adoptive transfer experiments, bmDCs were isolated from the femurs and tibias of hCD155-tg mice and cultured at 2.5×10^6^ cells/ml in RPMI + 10% FBS containing 300ng/ml human recombinant FLT3L (Thermo-Fisher #PHC9411) for 9 days. The non-adherent cells were treated with PVSRIPO or mock (24h), labeled with Celltrace Far Red (Thermo Fisher) and used for adoptive transfer directly into palpable E0771 tumors (∼1×10^6^ cells/mouse). CD11c^+^ (Biolegend CD11c-APC-Cy7 #117324) expression and Celltrace Far Red expression were checked by flow cytometry.

### Biodistribution Studies

CT2A or B16 tumor-bearing mice were treated with PVSRIPO as shown, euthanized, and blood was collected by exsanguination into EDTA-coated collection tubes (BD). Tissues (tumors, brain hemispheres, tdLN, spleen, kidney) were dissected and stored on ice in 1ml PBS. Tissue samples were weighed, tdLN and spleen were crushed with 1 ml PBS in a cell strainer, and the remaining tissues were homogenized using a drill. All tissues suspensions were brought to 1ml volume, subjected to 3 freeze-thaw cycles, and centrifugation (1000G) to remove large debris. The supernatant was collected for plaque assays as described previously.^18^

### Flow Cytometry Analyses

Cell and protein analyses were performed by flow cytometry. Multiple organs (tumor, brain, and tdLN) were harvested from euthanized mice and stored on ice in RPMI (Thermo Fisher). Tissues were sliced into small chunks with a disposable scalpel (Bard-Parker) and digested in 5ml of RPMI with DNase and Liberase (100μg/ml, each) (30min; 37°C, shaking). The digested material was then processed through a 0.7μm cell strainer (Genesee Scientific) and spun down at 500G (3min). Erythrocytes were removed with incubation in Red Blood Cell Lysing Buffer Hybri-Max (Sigma-Aldrich). Where required, cell subsets were isolated using various EasySep Selection kits (Stem Cell). EasySep Mouse Pan-DC Enrichment Kit II (#19863) and EasySep Mouse CD11c Positive Selection Kit II (#18780) were used to isolate DCs for further analysis. Isolation accuracy was verified by flow cytometry. Zombie-Aqua Fixable Viability dye (BioLegend) was used to stain dead cells in samples. All samples were resuspended in FACs buffer (2% FBS in PBS) containing 1% Mouse TruStain FcX FC Receptor Blocking solution (BioLegend) and incubated at 20°C (30min). Flow antibodies or isotype controls were then added to each tube; antibodies used in flow cytometry analyses are listed in Supplementary Table 3. Cells were incubated with the antibodies (1h, 20°C), with shaking every 15 min. Cells were then rinsed with 1ml PBS and spun down at 500G. After 30min, the cells were washed with FACs buffer (2ml) and pelleted. After a final wash, cells were resuspended in 300μl FACs buffer and analyzed by flow cytometry on Aurora (Cytek Biosciences) or LSRFortessa (BD) instruments. For intracellular vRNA staining, we used combined HCR-FISH (see below) with flow cytometry. For this procedure, processed cell suspensions were fixed in 4% paraformaldehyde (1h, 20°C). Fixed cells were pelleted by centrifugation for 3min at 500 G, washed 4 times with PBS-T (PBS, Tween-20 0.5%), and resuspended in 70% EtOH and stored (24h, 4°C). Cells were then washed with PBS-T and resuspended with probe hybridization buffer for HCR-FISH (Molecular Instruments Reagents; see below for detailed steps). Upon completion of all HCR-FISH steps the cells were washed and resuspended in FACS buffer for FACS analysis.

For human peripheral blood mononuclear cell (PBMC) flow cytometry analyses, two panels were used to define myeloid and T cell proportions and phenotypes. PBMCs were thawed at 37°C, washed in 7ml RPMI-10 (Gibco) + 10% FBS, resuspended in 1ml RPMI-10 containing 20μg of DNAse I (Roche), and incubated for 10min at 37°C. Cells were counted and diluted to 1×10^6^ cells/ml in RPMI-10, from which 300μl of the sample was used for PBMC stimulation assays. The remaining cell material was pelleted, resuspended in PBS (Gibco) containing 1:250 Zombie-NIR live/dead stain (BioLegend) and incubated for 15min (20°C), washed in FACs buffer (PBS containing 2% FBS), and resuspended in 100μl of FACs buffer containing 1:100 Human Tru-Stain FC block (BioLegend) for a minimum of 10min. Fifty microliters of cell suspension were transferred to panel-specific tubes to which 50μl of FACs buffer containing panel-specific, fluorophore-conjugated antibodies against the surface proteins in Supplementary Table 3 (1h, 20°C), followed by washing with 250μl of FACs buffer. PBMCs stained in panel 1 (surface stain only) were resuspended in 120μl FACs buffer and analyzed on a Fortessa X20 Flow Cytometer within 24h (BD Biosciences). PBMCs stained in panel 2 (T cells) were fixed and permeabilized using the FoxP3/Transcription factor staining buffer set (Thermo-Fisher) following manufacturer’s instructions and were stained with intracellular antibodies overnight at 4°C (using panel-specific antibodies; Supplementary Table 3). The next day PBMCs were washed in 1x Perm buffer (Thermo-Fisher) and resuspended in FACs buffer, followed by analysis on a Cytek Aurora within 72h (Cytek Biosciences). Each panel was validated for appropriate antibody titration and compensation (Fortessa X20)/unmixing (Cytek Aurora) using de-identified, leukopak-derived (Stemcell Technologies) PBMCs prior to commencing on-study sample analyses. Established negative cell populations, fluorescence minus one controls, and isotype controls were used to define positive vs negative staining, confirm appropriate compensation/spectral unmixing, and confirm specificity of antibody staining (see Supplementary Fig. 6d,e for representative gating). All flow cytometry data were analyzed using FlowJo v10.10 (BD Biosciences).

### RNA Analyses

Total cellular/tissue RNA was isolated from cell pellets or whole tissues (cLN and brain hemispheres) by resuspension in Trizol Reagent (Invitrogen) and frozen at -80°C. Chloroform (250μl) was added and mixed before the sample was spun at 12,000 rpm for (15min, 4°C) to induce phase separation. The aqueous layer was transferred to a new tube, mixed with isopropanol (500μl), and spun down at 12,000 rpm (10min, 4°C). The supernatant was removed, the RNA pellet was washed with 70% EtOH twice, dried on a 37°C heating block and finally resuspended in nuclease-free water. Column purification (Thermo Fisher) was then performed to concentrate and remove impurities. RNA concentration was checked using a nanodrop. Purified RNA was dried using a speedvac and sent to GeneWiz for bulk RNA sequencing. For RT-qPCR analysis, the RNA was treated with TURBO DNase (2U) (Thermo Fisher) and reverse-transcribed into cDNA using the High Capacity cDNA Reverse Transcription Kit (Thermo Fisher) following the manufacturer’s instructions. Two-step qPCR was performed with custom primer pairs for PVSRIPO^52^ and standard primer pairs for Ifit3 (Applied Biosystems Mm01704846_s1 FAM), Isg15 (*ibid*. Mm01705338_s1 FAM), Irf7 (*ibid*. Mm00516793_g1 FAM), and Gapdh (*ibid*. Mm99999915_g1 VIC) using TaqMan Fast Advanced Master Mix (Applied Biosystems).

### Microscopy

Ipsilateral (to the site of the glioma) deep cLN were harvested from mice bearing CT2A murine gliomas and treated with PVSRIPO/mock as shown. The tissues were embedded in OCT and flash frozen in liquid nitrogen, cut into 7μm cryosections, and mounted on slides. For immunofluorescence (IF) staining of CD11c (Cell Signaling #97585), MECA-79 (Biolegend #120802), and ISG15 (Invitrogen #703132) sections were fixed in 4% paraformaldehyde (20min), permeabilized (Triton X-100, 0.5%), incubated in blocking buffer (donkey serum, 5%), incubated in primary antibody (1:200 dilution; 24h, 4°C), rinsed (TBS, 3x), incubated with secondary anti-species antibody (donkey anti-rat; Invitrogen #A48272; 2h, 20°C), rinsed (TBS, 3x), and counterstained with 4′,6-diamidino-2-phenylindole (DAPI). Where indicated, HCR-FISH was performed on the stained slides for imaging of PVSRIPO (-)strand vRNA with specific probes (Molecular Instruments Reagents) (Supplementary Table 1). Slides were rinsed with PBS-T and incubated with probe hybridization buffer (Molecular Instruments Reagents) for pre-hybridization without probes (30min; 37°C). The probe solution was prepared by adding 2pM of each probe set to 100μl probe hybridization buffer (preheated to 37°C) and added directly onto the sample (24h, 37°C). After probe hybridization, the slides were washed with probe wash buffer (Molecular Instruments Reagents) to remove the probe solution. Hairpin amplification occurred by incubating the cells with snap-cooled h1 and h2 hairpins in amplification buffer (Molecular Instruments Reagents) (24h in the dark, 20°C). The hairpin solution was removed, the slides were rinsed five times with SSC buffer (150mM sodium chloride, 15mM sodium citrate) and mounted for imaging. Slides were imaged on a Leica Stellaris 8 confocal instrument in the Light Microscopy Core Facility, Duke University School of Medicine.

### Isolation and Analysis of Extracellular Vesicles

Extracellular vesicles were isolated from the supernatant of A375 melanoma cells as described previously.^42^ Briefly, A375 human melanoma cells cultured in serum-free DMEM were infected with PVSRIPO at a multiplicity of infection of 10. Two days after, the supernatant was collected and spun down at 1000G to remove cell debris. A high-speed centrifuge was then used to pellet vesicular material at 10,000 rpm and the resulting pellet was resuspended in PBS. An exosome isolation kit (Systems Biosciences) was used to affinity-purify vesicles via tetraspanins CD63, CD81, and CD9. Flow cytometry (CD63-BV786, CD81-PerCPCy5, CD9-PECy7, Biolegend) was used to confirm the presence of these vesicle markers. Plaque assays were used to determine the presence of intact viral particles in the isolated vesicles. Vesicles were imaged using dSTORM microscopy, using CellMaskRed (Thermo Fisher), A12 anti-poliovirus capsid antibody (1:1000), and AlexaFluor 568-CD63 antibody (R&D Systems).^59^ A nanoparticle tracker was used to measure the size and volume of vesicles.

### RNA Sequencing and Mapping

Total RNA sequencing was performed by Genewiz using PolyA selection for mRNA from samples with RIN score of ≥6. FASTQ files were evaluated using FASTQC/0.11.7 for low-quality reads and adapter content, which was removed using Trimmomatic/0.39 using adapters from Trimmomatic-0.35/adapters/TruSeq2-PE.fa and a sliding window of 4:30. Mapping was performed using STAR/2.7.5c. Reference index was generated from mouse reference genome GRCm39, Sabin reference genome GCF_008766755.1 (identical coding sequence to PVSRIPO), BSD (blasticidin resistance) from pLenti6.2 V5-DEST, luciferase from pHIV-Luciferase, and human PVR variant 1 NM_006505. FASTA and GTF files were combined sequentially to create a single FASTA and GTF file which was used to create the index. Trimmed FASTQ files were mapped to the index to produce count tables. Data analysis and visualization were performed using R version 4.4.0 (2024-04-24) and packages: purrr_1.0.2, dplyr_1.1.4, biomaRt_2.60.1, DESeq2_1.44.0, tibble_3.2.1, ggplot2_3.5.1, tidyr_1.3.1, reshape2_1.4.4, ggrepel_0.9.6, msigdbr_7.5.1, stringr_1.5.1, fgsea_1.30.0, org.Mm.eg.db_3.19.1, clusterProfiler_4.12.6, r2r_0.1.1, GO.db_3.19.1, data.tree_1.1.0, gridExtra_2.3, factoextra_1.0.7, and limma_3.60.6. Genes with <10 counts across all samples were dropped for analysis. Counts were normalized for analysis and further variance stabilization transformed for analysis and visualization by PCA. Outliers were visually identified by PCA analysis where samples were significant outliers from their treatment groups. For visualization, batch effects were removed using limma, whereas batches were included as a factor in DESeq2 for further analysis. Adjusted P values as calculated from DESeq2 are used for significance where gene expression was identified by RNAseq. Where genes were selected for being significantly differentially expressed, the adjusted p value was <0.05, and where genes were determined to be highly differentially expressed, they further had an absolute log2 fold change of at least 2 (4-fold). In cLN, analysis was performed only on protein coding genes as identified using Ensembl from the mmusculus_gene_ensembl dataset gene_biotype variable. GSEA pathways were accessed from MSigDB using msigdbr. Hallmark pathways (category “H”) were used to analyze data with fgsea. GO enrichment was performed using genes that were significantly differentially upregulated as determined by DESeq2 and using the enrichGO function from the clusterProfiler package. Gene ontologies were further simplified with a cutoff of 0.5.

Sequencing libraries of single nuclei from formalin-fixed, paraffin-embedded biopsy tissue were generated using the 10x Genomics flex gene expression kit on a 10x Chromium X workstation per manufacturer’s instructions. Cell Ranger (10x Genomics) v8.0.1 was used to demultiplex and align reads to Human genome version GRCh38-2020-A. Cell clusters were generated using Seurat’s FindClusters() function and visualized by Uniform Manifold Approximation and Projection (UMAP). Cell identities and T cell/immune cell phenotypes were determined using known markers for T cells (Supplementary Table 2).

### Statistics

R and GraphPad were used for statistical analyses. Specific statistical methods used, where applicable, are indicated in the relevant figure legends.

## References

1 Broz, M. L. et al. Dissecting the tumor myeloid compartment reveals rare activating antigen-presenting cells critical for T cell immunity. Cancer Cell 26, 638–652, doi:10.1016/j.ccell.2014.09.007 (2014).

2 Roberts, E. W. et al. Critical Role for CD103(+)/CD141(+) Dendritic Cells Bearing CCR7 for Tumor Antigen Trafficking and Priming of T Cell Immunity in Melanoma. Cancer Cell 30, 324–336, doi:10.1016/j.ccell.2016.06.003 (2016).

3 Hildner, K. et al. Batf3 deficiency reveals a critical role for CD8alpha+ dendritic cells in cytotoxic T cell immunity. Science 322, 1097–1100, doi:10.1126/science.1164206 (2008).

4 Spranger, S., Dai, D., Horton, B. & Gajewski, T. F. Tumor-Residing Batf3 Dendritic Cells Are Required for Effector T Cell Trafficking and Adoptive T Cell Therapy. Cancer Cell 31, 711–723 e714, doi:10.1016/j.ccell.2017.04.003 (2017).

5 Chen, D. S. & Mellman, I. Oncology meets immunology: the cancer-immunity cycle. Immunity 39, 1–10, doi:10.1016/j.immuni.2013.07.012 (2013).

6 Diamond, M. S. et al. Type I interferon is selectively required by dendritic cells for immune rejection of tumors. J Exp Med 208, 1989–2003, doi:10.1084/jem.20101158 (2011).

7 Sporri, R. & Reis e Sousa, C. Inflammatory mediators are insufficient for full dendritic cell activation and promote expansion of CD4+ T cell populations lacking helper function. Nat Immunol 6, 163–170, doi:10.1038/ni1162 (2005).

8 Freigang, S., Probst, H. C. & van den Broek, M. DC infection promotes antiviral CTL priming: the ’Winkelried’ strategy. Trends Immunol 26, 13–18, doi:10.1016/j.it.2004.11.013 (2005).

9 Joffre, O., Nolte, M. A., Sporri, R. & Reis e Sousa, C. Inflammatory signals in dendritic cell activation and the induction of adaptive immunity. Immunol Rev 227, 234–247, doi:10.1111/j.1600-065X.2008.00718.x (2009).

10 Trinchieri, G. Type I interferon: friend or foe? J Exp Med 207, 2053–2063, doi:10.1084/jem.20101664 (2010).

11 De Giovanni, M. et al. Spatiotemporal regulation of type I interferon expression determines the antiviral polarization of CD4(+) T cells. Nat Immunol 21, 321–330, doi:10.1038/s41590-020-0596-6 (2020).

12 Wang, Y. et al. Timing and magnitude of type I interferon responses by distinct sensors impact CD8 T cell exhaustion and chronic viral infection. Cell Host Microbe 11, 631–642, doi:10.1016/j.chom.2012.05.003 (2012).

13 Izaguirre, A. et al. Comparative analysis of IRF and IFN-alpha expression in human plasmacytoid and monocyte-derived dendritic cells. J Leukoc Biol 74, 1125–1138, doi:10.1189/jlb.0603255 (2003).

14 Bodian, D. Emerging concept of poliomyelitis infection. Science 122, 105–108 (1955).

15 Sabin, A. B. Pathogenesis of poliomyelitis; reappraisal in the light of new data. Science 123, 1151–1157 (1956).

16 Burrows, M. T. Is poliomyelitis a disease of the lymphatic system? Arch. Int. Med. 48, 33–50 (1931).

17 Shen, L. et al. Pathogenic Events in a Nonhuman Primate Model of Oral Poliovirus Infection Leading to Paralytic Poliomyelitis. J Virol 91, doi:10.1128/JVI.02310-16 (2017).

18 Gromeier, M., Alexander, L. & Wimmer, E. Internal ribosomal entry site substitution eliminates neurovirulence in intergeneric poliovirus recombinants. Proc Natl Acad Sci U S A 93, 2370–2375 (1996).

19 Dobrikov, M. I. et al. Early enterovirus translation deficits extend viral RNA replication and elicit sustained MDA5-directed innate signaling. mBio 14, e0191523, doi:10.1128/mbio.01915-23 (2023).

20 Dobrikov, M. I. et al. PKR Binds Enterovirus IRESs, Displaces Host Translation Factors, and Impairs Viral Translation to Enable Innate Antiviral Signaling. mBio 13, e0085422, doi:10.1128/mbio.00854-22 (2022).

21 Hsu, N. Y. et al. Viral reorganization of the secretory pathway generates distinct organelles for RNA replication. Cell 141, 799–811, doi:10.1016/j.cell.2010.03.050 (2010).

22 Brown, M. C. et al. Viral infection of cells within the tumor microenvironment mediates antitumor immunotherapy via selective TBK1-IRF3 signaling. Nat Commun 12, 1858, doi:10.1038/s41467-021-22088-1 (2021).

23 Kato, H. et al. Differential roles of MDA5 and RIG-I helicases in the recognition of RNA viruses. Nature 441, 101–105, doi:10.1038/nature04734 (2006).

24 Brown, M. C. et al. Cancer immunotherapy with recombinant poliovirus induces IFN-dominant activation of dendritic cells and tumor antigen-specific CTLs. Sci Transl Med 9, doi:10.1126/scitranslmed.aan4220 (2017).

25 Mosaheb, M. M. et al. Genetically stable poliovirus vectors activate dendritic cells and prime antitumor CD8 T cell immunity. Nat Commun 11, 524, doi:10.1038/s41467-019-13939-z (2020).

26 Koike, S. et al. Transgenic mice susceptible to poliovirus. Proc Natl Acad Sci U S A 88, 951–955 (1991).

27 Yang, Y. et al. Polio virotherapy targets the malignant glioma myeloid infiltrate with diffuse microglia activation engulfing the CNS. Neuro Oncol 25, 1631–1643, doi:10.1093/neuonc/noad052 (2023).

28 Rustenhoven, J. & Kipnis, J. Brain borders at the central stage of neuroimmunology. Nature 612, 417–429, doi:10.1038/s41586-022-05474-7 (2022).

29 Ren, R. & Racaniello, V. R. Human poliovirus receptor gene expression and poliovirus tissue tropism in transgenic mice. J Virol 66, 296–304, doi:10.1128/JVI.66.1.296-304.1992 (1992).

30 Gromeier, M., Lachmann, S., Rosenfeld, M. R., Gutin, P. H. & Wimmer, E. Intergeneric poliovirus recombinants for the treatment of malignant glioma. Proc Natl Acad Sci U S A 97, 6803–6808 (2000).

31 Porritt, R. A. & Hertzog, P. J. Dynamic control of type I IFN signalling by an integrated network of negative regulators. Trends Immunol 36, 150–160, doi:10.1016/j.it.2015.02.002 (2015).

32 Forster, R. et al. CCR7 coordinates the primary immune response by establishing functional microenvironments in secondary lymphoid organs. Cell 99, 23–33, doi:10.1016/s0092-8674(00)80059-8 (1999).

33 Louveau, A. et al. Structural and functional features of central nervous system lymphatic vessels. Nature 523, 337–341, doi:10.1038/nature14432 (2015).

34 Kim, M. W. et al. Endogenous self-peptides guard immune privilege of the central nervous system. Nature 637, 176–183, doi:10.1038/s41586-024-08279-y (2025).

35 Bowman-Kirigin, J. A. et al. The Conventional Dendritic Cell 1 Subset Primes CD8+ T Cells and Traffics Tumor Antigen to Drive Antitumor Immunity in the Brain. Cancer Immunol Res 11, 20–37, doi:10.1158/2326-6066.CIR-22-0098 (2023).

36 Johanns, T. M. et al. Endogenous Neoantigen-Specific CD8 T Cells Identified in Two Glioblastoma Models Using a Cancer Immunogenomics Approach. Cancer Immunol Res 4, 1007–1015, doi:10.1158/2326-6066.CIR-16-0156 (2016).

37 De Stefani, A. et al. Improved survival with perilymphatic interleukin 2 in patients with resectable squamous cell carcinoma of the oral cavity and oropharynx. Cancer 95, 90–97, doi:10.1002/cncr.10654 (2002).

38 Cortesina, G. et al. Treatment of recurrent squamous cell carcinoma of the head and neck with low doses of interleukin-2 injected perilymphatically. Cancer 62, 2482–2485, doi:10.1002/1097-0142(19881215)62:12<2482::aid-cncr2820621205>3.0.co;2-9 (1988).

39 Girard, J. P., Moussion, C. & Forster, R. HEVs, lymphatics and homeostatic immune cell trafficking in lymph nodes. Nat Rev Immunol 12, 762–773, doi:10.1038/nri3298 (2012).

40 Blanchard, L. & Girard, J. P. High endothelial venules (HEVs) in immunity, inflammation and cancer. Angiogenesis 24, 719–753, doi:10.1007/s10456-021-09792-8 (2021).

41 Wendland, M. et al. Lymph node T cell homeostasis relies on steady state homing of dendritic cells. Immunity 35, 945–957, doi:10.1016/j.immuni.2011.10.017 (2011).

42 Chen, Y. H. et al. Phosphatidylserine vesicles enable efficient en bloc transmission of enteroviruses. Cell 160, 619–630, doi:10.1016/j.cell.2015.01.032 (2015).

43 Harrell, M. I., Iritani, B. M. & Ruddell, A. Lymph node mapping in the mouse. J Immunol Methods 332, 170–174, doi:10.1016/j.jim.2007.11.012 (2008).

44 Cabeza-Cabrerizo, M., Cardoso, A., Minutti, C. M., Pereira da Costa, M. & Reis e Sousa, C. Dendritic Cells Revisited. Annu Rev Immunol 39, 131–166, doi:10.1146/annurev-immunol-061020-053707 (2021).

45 Alcami, A. & Koszinowski, U. H. Viral mechanisms of immune evasion. Trends Microbiol 8, 410–418, doi:10.1016/s0966-842x(00)01830-8 (2000).

46 Goubau, D., Deddouche, S. & Reis e Sousa, C. Cytosolic sensing of viruses. Immunity 38, 855–869, doi:10.1016/j.immuni.2013.05.007 (2013).

47 Hansen, T. H. & Bouvier, M. MHC class I antigen presentation: learning from viral evasion strategies. Nat Rev Immunol 9, 503–513, doi:10.1038/nri2575 (2009).

48 Levy, D. E. & Garcia-Sastre, A. The virus battles: IFN induction of the antiviral state and mechanisms of viral evasion. Cytokine Growth Factor Rev 12, 143–156, doi:10.1016/s1359-6101(00)00027-7 (2001).

49 Petersen, J. L., Morris, C. R. & Solheim, J. C. Virus evasion of MHC class I molecule presentation. J Immunol 171, 4473–4478 (2003).

50 Ploegh, H. L. Viral strategies of immune evasion. Science 280, 248–253 (1998).

51 Etchison, D., Milburn, S. C., Edery, I., Sonenberg, N. & Hershey, J. W. Inhibition of HeLa cell protein synthesis following poliovirus infection correlates with the proteolysis of a 220,000-dalton polypeptide associated with eucaryotic initiation factor 3 and a cap binding protein complex. J Biol Chem 257, 14806–14810 (1982).

52 Dobrikova, E. Y. et al. Attenuation of neurovirulence, biodistribution, and shedding of a poliovirus:rhinovirus chimera after intrathalamic inoculation in Macaca fascicularis. J Virol 86, 2750–2759, doi:10.1128/JVI.06427-11 (2012).

53 Desjardins, A. et al. Recurrent Glioblastoma Treated with Recombinant Poliovirus. N Engl J Med 379, 150–161, doi:10.1056/NEJMoa1716435 (2018).

54 Friedman, G. K. et al. Oncolytic HSV-1 G207 Immunovirotherapy for Pediatric High-Grade Gliomas. N Engl J Med 384, 1613–1622, doi:10.1056/NEJMoa2024947 (2021).

55 Lang, F. F. et al. Phase I Study of DNX-2401 (Delta-24-RGD) Oncolytic Adenovirus: Replication and Immunotherapeutic Effects in Recurrent Malignant Glioma. J Clin Oncol 36, 1419–1427, doi:10.1200/JCO.2017.75.8219 (2018).

56 Ling, A. L. et al. Clinical trial links oncolytic immunoactivation to survival in glioblastoma. Nature 623, 157–166, doi:10.1038/s41586-023-06623-2 (2023).

57 Todo, T. et al. Intratumoral oncolytic herpes virus G47Δ for residual or recurrent glioblastoma: a phase 2 trial. Nat Med 28, 1630–1639, doi:10.1038/s41591-022-01897-x (2022).

58 Brown, M. C. et al. Intratumor childhood vaccine-specific CD4(+) T-cell recall coordinates antitumor CD8(+) T cells and eosinophils. J Immunother Cancer 11, doi:10.1136/jitc-2022-006463 (2023).

59 Chambers, M. G., McNamara, R. P. & Dittmer, D. P. Direct Stochastic Optical Reconstruction Microscopy of Extracellular Vesicles in Three Dimensions. J Vis Exp, doi:10.3791/62845 (2021).

