## Supplemental Information for "Lymphotropic Virotherapy Engages DC and High Endothelial Venule Inflammation to Mediate Cancer *In Situ* Vaccination"

Andrea L. Ludwig, *et al*.

**Supplementary Table 1.** FISH probe sets (extended data relating to Figures 2c, 4)**.**

Probe set for detection of (+)strand vRNA of PVSRIPO:

5′-tagtaacgcggcttcgaaacaggacttctctcaagacccttccaagttcacc-3′
5′-gcccatcaaggatgtcctgataaaaacatccccaatgctaaactcgccaaac-3′
5′-agaggcttgcgggtatagcgatagagtactgcaattaacactgggaaactcc-3′
5′-tataaccacacaggaggcggctaattcagtagtcgcttatgggcgttggcct-3′
5′-atatctgagggacagcgaagccaatccagtggaccagccgacagaaccagac-3′
5′-ggcaggatccctgaagttcacgtttctgttctgtggatccatgatggcaact-3′
5′-caaactgttggtgtcatacgcgcctcctggagccgacccaccaaagaagcgt-3′
5′-ggaggcgatgttgggaacacatgtgatctgggacataggactgcagtcctca-3′
5′-tactatggtagtgccatggattagcaacaccacgtatcggcaaaccatagat-3′
5′-tagtttcaccgaaggcggatacatcagcgtcttctaccaaaccagaatagtc-3′
5′-gtccacgtactcgctacccccggatccatcacacttcgacggatacaaacaa-3′
5′-gggagtggtgattatggacgacctgaatcaaaacccagatggtgcggacatg-3′
5′-gctgttctgtcagatggtatcaacagtggagtttataccacccatggcatcc-3′
5′-ggaggagaaaggaatcctgtttacttcaaattacgttctagcatccacgaac-3′
5′-aagcagaatttcccccccactgtggcacacagtgatgcattagccaggcgc-3′
5′-caatatgtatgttcctgtcggtgctgtgactgaacagggatatctaaatctc-3′
5′-tgggcgccaaactgctcgtactctaatgtacaactttccaaccagagcagga-3′
5′-gtgtggtggagtcatcacatgtactgggaaagtcatcgggatgcatgttggt-3′
5′-gaacggttcacacgggtttgcagcggccctgaagcgatcatacttcactcag-3′
5′-tcaaggtgaaatccagtggatgagaccttcgaaggaagtgggatatccaatc-3′

Probe set for detection of (-)strand vRNA of PVSRIPO:

5′-ctgtacatgcacggtgtacccggacctacctaggtagtggtagtacatattt-3′
5′-ctctggtacggcgaataccccctagtgccccctggtggaatttggaggcgtta-3′
5′-attttgatagctggtgtgcatggtagtggtgttgctatccccggccagacac-3′
5′-atttccaaagaggtaatccaccgggcagaacctacgggcaggtgatgtctgg-3′
5′-ccgtaggtttattatctggtgcgggaacacaaaggcattccccaataacgtg-3′
5′-cgctttttgctctatatgtgtggtatctcgcatcaagcgcacgctgaagtca-3′
5′-actggcttcagtgtttgggagagcgtctctagacgttgccgcccccaccgtt-3′
5′-ggccccagtttccactgcggtgagtgccggaatttccttggagtgtgctggt-3′
5′-atgttgtacaacatgtctggtttgcactgtatcagaagggactagtggattt-3′
5′-aaatagcttatccttattcttggtggaagctgagttatccacggttataatg-3′
5′-tggtccttggaacaaagcctccatacaattgccaatgttggatcttctgttt-3′
5′-acattcaggaggggactcgtcttgatgtcaatcttcaagtctttatactgg-3′
5′-ctcacagtaatctctcacctcctgggagtcaactgcttggagcaagtcattg-3′
5′-gatgttcctttctgtttgaacctggctggtgatgttgactatccaacccttc-3′
5′-tccagccactgcggcgaaggttgtcaccgcttgtagaattgtcattgccctg-3′
5′-cacttcagtaattttgttacccacgtacttggagaaaattgcctcctcaaag-3′
5′-accatcagtgccatacatggcatcctccaagcacatttgttctgtgttgatg-3′
5′-tcccattgctacataagggtagccagcactggtggacaaatcaagtgcttct-3′
5′-atcctttacataagtcaccagtgggaggttgattccatatgtgtcgagcagt-3′
5′-agcatataggttcccaaaagccattctcattgccactgagtcattcaaacta-3′

**Supplementary Table 2.** Top-20 differentially expressed genes in tumor-infiltrating T cell subsets (extended data relating to Figure 5d).

**a** activated CD4 subset (aCD4)

| gene | pct.1 | pct.2 | avg_log2FC | p_val | p_val_adj |
|---|---|---|---|---|---|
| CD40LG | 0.349 | 0.069 | 2.4301058 | 5.221660e−27 | 6.720277e−23 |
| CD4 | 0.559 | 0.231 | 1.4198071 | 1.236034e−22 | 1.590776e−18 |
| SAMD9L | 0.724 | 0.396 | 1.2034189 | 2.104773e−21 | 2.708843e−17 |
| IFI44 | 0.691 | 0.440 | 1.3759772 | 2.120355e−19 | 2.728896e−15 |
| IFI44L | 0.523 | 0.256 | 1.8038932 | 1.744340e−17 | 2.244966e−13 |
| TYMP | 0.507 | 0.239 | 1.3235589 | 2.136258e−16 | 2.749364e−12 |
| SLAMF1 | 0.641 | 0.363 | 0.9382421 | 1.371095e−15 | 1.764599e−11 |
| SEC61G | 0.408 | 0.171 | 1.4443537 | 2.771670e−15 | 3.567139e−11 |
| C3 | 0.586 | 0.303 | 0.8018490 | 6.091458e−15 | 7.839707e−11 |
| EPSTI1 | 0.457 | 0.221 | 1.1825250 | 4.672900e−14 | 6.014022e−10 |
| CTSB | 0.579 | 0.333 | 1.1155339 | 1.258003e−13 | 1.619049e−09 |
| DDX60 | 0.368 | 0.159 | 1.2409713 | 1.768020e−12 | 2.275442e−08 |
| ADAM19 | 0.549 | 0.309 | 0.9682334 | 1.904402e−12 | 2.450965e−08 |
| STAT1 | 0.641 | 0.477 | 1.1504910 | 3.834800e−12 | 4.935387e−08 |
| ST8SIA1 | 0.454 | 0.231 | 1.0858623 | 8.472863e−12 | 1.090457e−07 |
| IFI35 | 0.336 | 0.145 | 1.3653486 | 1.190644e−11 | 1.532359e−07 |
| UBA7 | 0.546 | 0.306 | 0.8360013 | 2.037744e−11 | 2.622577e−07 |
| PARP14 | 0.599 | 0.368 | 0.8678754 | 2.742203e−11 | 3.529215e−07 |
| CD226 | 0.342 | 0.147 | 1.2152618 | 3.208202e−11 | 4.128956e−07 |
| SERINC5 | 0.352 | 0.157 | 1.1886188 | 3.245539e−11 | 4.177008e−07 |

**b** activated CD8 subset (aCD8)

| gene | pct.1 | pct.2 | avg_log2FC | p_val | p_val_adj |
|---|---|---|---|---|---|
| CD8A | 0.770 | 0.182 | 2.6364417 | 1.180370e−61 | 1.519136e−57 |
| CTSW | 0.765 | 0.222 | 2.0294290 | 1.068624e−48 | 1.375319e−44 |
| NKG7 | 0.750 | 0.202 | 1.8933024 | 1.158123e−47 | 1.490504e−43 |
| CCL5 | 0.877 | 0.466 | 2.1178782 | 1.295030e−46 | 1.666704e−42 |
| SLAMF7 | 0.623 | 0.142 | 2.2115217 | 8.350876e−44 | 1.074758e−39 |
| KLRK1 | 0.814 | 0.266 | 1.5122761 | 1.191619e−41 | 1.533614e−37 |
| GZMK | 0.681 | 0.262 | 1.8817937 | 2.169609e−32 | 2.792286e−28 |
| GZMA | 0.848 | 0.521 | 1.2040657 | 4.643745e−25 | 5.976500e−21 |
| ITGA1 | 0.480 | 0.143 | 1.9561780 | 6.841243e−25 | 8.804680e−21 |
| EPHA1 | 0.525 | 0.169 | 1.7300349 | 1.716373e−24 | 2.208972e−20 |
| TRGC2 | 0.461 | 0.135 | 1.7654979 | 6.768986e−24 | 8.711685e−20 |
| ZNF683 | 0.314 | 0.060 | 2.8879994 | 1.259810e−23 | 1.621375e−19 |
| GZMH | 0.417 | 0.117 | 2.3465642 | 1.517811e−23 | 1.953422e−19 |
| MATK | 0.515 | 0.169 | 1.6275536 | 2.877642e−23 | 3.703526e−19 |
| EOMES | 0.466 | 0.148 | 1.6966554 | 6.096963e−22 | 7.846791e−18 |
| TRGC1 | 0.304 | 0.066 | 2.3114113 | 2.645854e−20 | 3.405215e−16 |
| MT−ATP6 | 0.936 | 0.844 | 0.8663034 | 5.036148e−20 | 6.481523e−16 |
| NELL2 | 0.431 | 0.139 | 1.7823115 | 1.169391e−19 | 1.505006e−15 |
| KLRC4 | 0.456 | 0.138 | 0.8832991 | 1.734093e−19 | 2.231778e−15 |
| CCL4 | 0.373 | 0.103 | 1.5837151 | 2.240516e−19 | 2.883544e−15 |

**c** inactive CD4 subset (iaCD4)

| gene | pct.1 | pct.2 | avg_log2FC | p_val | p_val_adj |
|---|---|---|---|---|---|
| HSPA1B | 0.593 | 0.085 | 4.148888 | 6.773937e−67 | 8.718057e−63 |
| HSPA1A | 0.490 | 0.075 | 3.725893 | 1.860220e−50 | 2.394104e−46 |
| DNAJB1 | 0.544 | 0.156 | 2.889905 | 5.537111e−40 | 7.126261e−36 |
| HSPB1 | 0.475 | 0.135 | 3.006246 | 1.278192e−34 | 1.645033e−30 |
| CXCR4 | 0.700 | 0.379 | 1.854868 | 6.483817e−33 | 8.344673e−29 |
| NR4A1 | 0.270 | 0.034 | 3.427595 | 1.032185e−26 | 1.328422e−22 |
| VEGFA | 0.624 | 0.352 | 1.974187 | 2.818857e−25 | 3.627869e−21 |
| MT−ND6 | 0.886 | 0.665 | 1.183797 | 5.902095e−22 | 7.595996e−18 |
| TSC22D3 | 0.411 | 0.139 | 2.191116 | 7.610487e−22 | 9.794697e−18 |
| CCN1 | 0.217 | 0.034 | 3.648124 | 2.224903e−19 | 2.863450e−15 |
| TXNIP | 0.867 | 0.678 | 1.140115 | 3.789272e−19 | 4.876793e−15 |
| EGR1 | 0.395 | 0.153 | 1.962023 | 4.896272e−19 | 6.301502e−15 |
| FOS | 0.471 | 0.239 | 1.748702 | 3.135576e−17 | 4.035486e−13 |
| HBA2 | 0.228 | 0.049 | 2.385090 | 1.295727e−16 | 1.667600e−12 |
| RYR3 | 0.221 | 0.045 | 2.686010 | 2.307560e−16 | 2.969829e−12 |
| FKBP5 | 0.346 | 0.141 | 2.035641 | 5.223131e−15 | 6.722169e−11 |
| CD163 | 0.221 | 0.052 | 2.691949 | 5.620168e−15 | 7.233157e−11 |
| FOSB | 0.160 | 0.022 | 2.804315 | 5.856907e−15 | 7.537839e−11 |
| DTNA | 0.502 | 0.321 | 1.562360 | 1.049533e−13 | 1.350749e−09 |
| DUSP1 | 0.300 | 0.117 | 2.105182 | 5.588147e−13 | 7.191945e−09 |

**d** NK subset (NK)

| gene | pct.1 | pct.2 | avg_log2FC | p_val | p_val_adj |
|---|---|---|---|---|---|
| KLRD1 | 0.935 | 0.042 | 4.983440 | 3.917621e−99 | 5.041979e−95 |
| NCR1 | 0.391 | 0.006 | 6.305368 | 1.273076e−58 | 1.638449e−54 |
| GNLY | 0.674 | 0.046 | 5.163927 | 1.630089e−56 | 2.097924e−52 |
| SH2D1B | 0.261 | 0.004 | 6.610222 | 2.086003e−40 | 2.684686e−36 |
| FGR | 0.457 | 0.027 | 4.668566 | 2.300863e−40 | 2.961210e−36 |
| KLRF1 | 0.304 | 0.008 | 5.758473 | 1.003977e−38 | 1.292119e−34 |
| TRDC | 0.435 | 0.028 | 3.110102 | 2.987553e−35 | 3.844980e−31 |
| KLRC3 | 0.435 | 0.032 | 4.632943 | 3.336073e−34 | 4.293525e−30 |
| KLRC1 | 0.435 | 0.035 | 3.821976 | 3.836245e−31 | 4.937247e−27 |
| NMUR1 | 0.348 | 0.028 | 3.732478 | 8.903892e−25 | 1.145931e−20 |
| FGFBP2 | 0.152 | 0.002 | 6.217905 | 2.275780e−23 | 2.928928e−19 |
| TXK | 0.478 | 0.064 | 3.217905 | 5.259365e−23 | 6.768803e−19 |
| OSBPL5 | 0.283 | 0.020 | 4.217905 | 2.706902e−22 | 3.483784e−18 |
| NKG7 | 0.826 | 0.299 | 2.394620 | 1.351386e−17 | 1.739234e−13 |
| S1PR5 | 0.217 | 0.015 | 4.465833 | 1.582174e−17 | 2.036258e−13 |
| KLRC4 | 0.652 | 0.186 | 2.565828 | 4.261221e−17 | 5.484192e−13 |
| GZMB | 0.261 | 0.029 | 3.839393 | 7.705999e−15 | 9.917621e−11 |
| SPON2 | 0.435 | 0.090 | 3.151410 | 1.272004e−14 | 1.637069e−10 |
| AOAH | 0.783 | 0.348 | 2.189523 | 1.336717e−14 | 1.720355e−10 |
| MYOM2 | 0.087 | 0.001 | 5.539833 | 2.389566e−14 | 3.075372e−10 |

**e** Treg subset (Treg)

| gene | pct.1 | pct.2 | avg_log2FC | p_val | p_val_adj |
|---|---|---|---|---|---|
| FOXP3 | 0.718 | 0.013 | 6.257849 | 1.034124e−115 | 1.330917e−111 |
| LAYN | 0.388 | 0.007 | 6.264801 | 6.349957e−61 | 8.172395e−57 |
| CTLA4 | 0.659 | 0.078 | 3.801401 | 5.120860e−54 | 6.590547e−50 |
| TNFRSF18 | 0.647 | 0.092 | 4.084229 | 9.274042e−48 | 1.193569e−43 |
| RTKN2 | 0.482 | 0.040 | 4.599786 | 3.848333e−47 | 4.952804e−43 |
| TNFRSF4 | 0.435 | 0.035 | 4.645891 | 1.675026e−42 | 2.155759e−38 |
| IL2RA | 0.365 | 0.021 | 5.042409 | 1.257863e−41 | 1.618870e−37 |
| F5 | 0.329 | 0.015 | 5.023793 | 2.345662e−41 | 3.018867e−37 |
| CCR8 | 0.224 | 0.002 | 6.487194 | 7.341515e−38 | 9.448530e−34 |
| IKZF2 | 0.600 | 0.102 | 3.112798 | 4.329414e−37 | 5.571956e−33 |
| STAM | 0.624 | 0.136 | 2.728300 | 4.842475e−31 | 6.232265e−27 |
| IL1RL1 | 0.176 | 0.002 | 6.434726 | 2.830157e−29 | 3.642412e−25 |
| TNFRSF9 | 0.400 | 0.054 | 3.759566 | 2.839141e−28 | 3.653974e−24 |
| WHRN | 0.518 | 0.110 | 3.090772 | 7.874582e−26 | 1.013459e−21 |
| TIGIT | 0.765 | 0.280 | 2.513657 | 9.114927e−26 | 1.173091e−21 |
| TNFRSF1B | 0.812 | 0.327 | 2.081004 | 2.295075e−25 | 2.953762e−21 |
| IL1R1 | 0.153 | 0.002 | 5.767302 | 5.431795e−25 | 6.990720e−21 |
| IL12RB2 | 0.365 | 0.051 | 3.400153 | 1.399196e−24 | 1.800765e−20 |
| TBC1D4 | 0.529 | 0.125 | 2.611126 | 1.727989e−23 | 2.223921e−19 |
| PTGIR | 0.118 | 0.000 | 7.171692 | 6.722157e−23 | 8.651417e−19 |

**Supplementary Table 3.** Antibodies used in flow-cytometry analyses.

| Antigen | Fluorophore | Vendor | Cat # | Dilution | RRID |
|---|---|---|---|---|---|
| Mouse: | | | | | |
| CD45 | BUV395 | BD | 564279 | 100x | AB_2651134 |
| CD19 | PE | Biolegend | 152408 | 100x | AB_2629817 |
| CD3 | PE | Biolegend | 100206 | 100x | AB_312663 |
| NK1.1 | FITC | Biolegend | 108706 | 100x | AB_313393 |
| IA/IE | BV785 | Biolegend | 107645 | 100x | AB_2565977 |
| CD11c | APC | Biolegend | 117310 | 100x | AB_313779 |
| CD11c | APC/Cy7 | Biolegend | 117324 | 100x | AB_830649 |
| CD19 | BUV615 | BD | 751213 | 100x | AB_2875235 |
| CD3 | BUV496 | BD | 741117 | 100x | AB_2870707 |
| NK1.1 | BV480 | BD | 746265 | 100x | AB_2743597 |
| CD11c | BV421 | Biolegend | 117330 | 100x | AB_11219593 |
| F4/80 | BUV805 | BD | 749282 | 100x | AB_2873657 |
| IA/IE | BV711 | Biolegend | 107643 | 100x | AB_2565976 |
| CD3 | BV510 | Biolegend | 100233 | 100x | AB_2561387 |
| CD19 | BV510 | Biolegend | 115546 | 100x | AB_2562137 |
| NK1.1 | BV510 | Biolegend | 108738 | 100x | AB_2562217 |
| F4/80 | BV605 | Biolegend | 123133 | 100x | AB_2562305 |
| CD14 | BV421 | Biolegend | 123329 | 100x | AB_2721526 |
| CD11b | BV711 | Biolegend | 101236 | 100x | AB_11203704 |
| Ly6C | PerCP-Cy5.5 | BD | 560525 | 100x | AB_1727558 |
| F4/80 | PE-Cy5 | Biolegend | 123112 | 100x | AB_893482 |
| Ly6G | BV605 | Biolegend | 127639 | 100x | AB_2565880 |
| Siglec-F | PE-Cy7 | Biolegend | 155528 | 100x | AB_2890715 |
| CD3 | BV605 | Biolegend | 317322 | 100x | AB_2561911 |
| CD4 | FITC | Biolegend | 100406 | 100x | AB_312691 |
| CD8 | BV421 | Biolegend | 100738 | 100x | AB_11204079 |
| FoxP3 | PE-Cy7 | Invitrogen | 25-5773-82 | 100x | AB_891552 |
| GzmB | PE-Cy7 | Invitrogen | 25-8898-82 | 100x | AB_10853339 |
| Human: | | | | | |
| CD63 | BV786 | Biolegend | 353044 | 100x | AB_2814289 |
| CD81 | PerCP-Cy5 | Biolegend | 349508 | 100x | AB_2564019 |
| CD9 | PECy7 | Biolegend | 124816 | 100x | AB_2783075 |

| Antigen | Fluorophore | Vendor | Cat # | Dilution | RRID |
|---|---|---|---|---|---|
| Human: | | | | | |
| CD3 | BUV737 | BD | 612750 | 200x | AB_2870081 |
| CD4 | BV510 | Biolegend | 357420 | 200x | AB_2715940 |
| CD8 | BUV395 | BD | 563795 | 200x | AB_2722501 |
| CD127 | PEcy5 | Biolegend | 351324 | 200x | AB_10915554 |
| CD27 | BV650 | Biolegend | 302828 | 200x | AB_2562096 |
| CD25 | PerCP-Cy5.5 | Biolegend | 302626 | 200x | AB_2125478 |
| CD56 | AF700 | Biolegend | 362522 | 200x | AB_2564099 |
| gdTCR | BUV563 | BD | 748534 | 200x | AB_2872945 |
| OX40 | BUV496 | BD | 750646 | 200x | AB_2874774 |
| CD28 | BUV805 | BD | 748474 | 200x | AB_2872889 |
| ICOS | BV480 | BD | 566085 | 200x | AB_3674209 |
| IFNg | PE | Biolegend | 383304 | 100x | AB_2924583 |
| GzmB | PEcy7 | Biolegend | 372214 | 100x | AB_2728381 |
| CD62L | APC-Fire810 | Biolegend | 304866 | 200x | AB_2894453 |
| CCR7 | BV750 | Biolegend | 353254 | 200x | AB_2800945 |
| CD45RA | APC | Biolegend | 304112 | 200x | AB_314416 |
| CD45RO | BV786 | Biolegend | 304234 | 200x | AB_2563819 |
| CD39 | PEDazzle594 | Biolegend | 328224 | 200x | AB_2454319 |
| TIM3 | PACblu | Biolegend | 345042 | 200x | AB_2632854 |
| TCF1 | BV421 | BD | 566692 | 50x | AB_2869822 |
| TIGIT | BV605 | Biolegend | 372712 | 200x | AB_2632927 |
| LAG3 | AF488 | Biolegend | 369326 | 200x | AB_2721362 |
| PD1 | BV711 | Biolegend | 367428 | 200x | AB_2721555 |
| BTLA | BUV615 | BD | 751542 | 200x | AB_2875537 |
| CD3 | FITC | Biolegend | 300306 | 200x | AB_314042 |
| CD19 | BV711 | Biolegend | 302246 | 200x | AB_2562065 |
| CD14 | BV421 | Biolegend | 325628 | 200x | AB_2563296 |
| CD16 | PerCP-Cy5.5 | Biolegend | 302028 | 200x | AB_893262 |
| CD11b | APC | Biolegend | 982604 | 200x | AB_2632619 |
| PD-L1 | BV605 | Biolegend | 329724 | 200x | AB_2565926 |
| CD11c | BUV395 | BD | 563787 | 200x | AB_2744274 |
| CD141 | PEcy7 | Biolegend | 344110 | 200x | AB_2561623 |
| CD123 | BV510 | Biolegend | 306022 | 200x | AB_2562068 |
| CD1c | PECF594 | Biolegend | 331532 | 200x | AB_2565293 |
| CD1a | PECY5 | Biolegend | 300108 | 200x | AB_314022 |
| CD155 | PE | Biolegend | 337610 | 200x | AB_2174019 |
| HLADR | BV786 | Biolegend | 307642 | 200x | AB_2563461 |

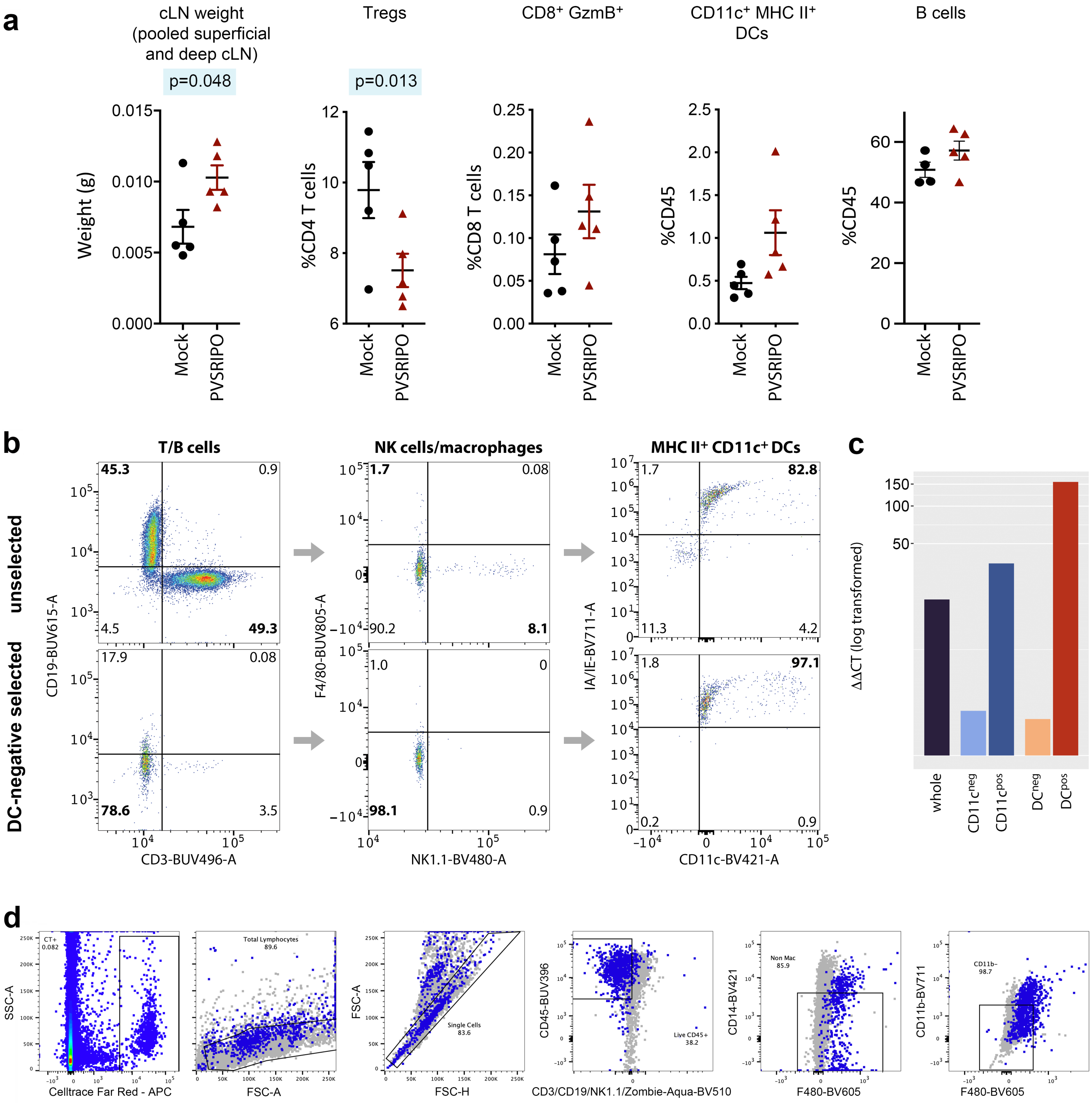
a
cLN weight (pooled superficial and deep cLN)
p=0.048
Weight (g)
0.000
0.005
0.010
0.015
Tregs
p=0.013
%CD4 T cells
6
8
10
12
CD8+ GzmB+
%CD8 T cells
0.00
0.05
0.10
0.15
0.20
0.25
CD11c+ MHC II+ DCs
%CD45
0.0
0.5
1.0
1.5
2.0
2.5
B cells
%CD45
0
20
40
60
Mock
PVSRIPO
b
T/B cells
NK cells/macrophages
MHC II+ CD11c+ DCs
unselected
DC-negative selected
45.3
0.9
4.5
49.3
17.9
0.08
78.6
3.5
1.7
0.08
90.2
8.1
1.0
0
98.1
0.9
1.7
82.8
11.3
4.2
1.8
97.1
0.2
0.9
CD19-BUV615-A
CD3-BUV496-A
F4/80-BUV805-A
NK1.1-BV480-A
IA/IE-BV711-A
CD11c-BV421-A
c
ΔΔCT (log transformed)
150
100
50
whole
CD11cneg
CD11cpos
DCneg
DCpos
d
CT+
0.082
SSC-A
Celltrace Far Red - APC
Total Lymphocytes
89.6
FSC-A
Single Cells
83.6
FSC-H
CD45-BUV396
Live CD45+
38.2
CD3/CD19/NK1.1/Zombie-Aqua-BV510
Non Mac
85.9
CD14-BV421
F480-BV605
CD11b-
98.7
CD11b-BV711
F480-BV605

**Supplementary Fig. 1.** Extended data on cLN phenotypes after IT PVSRIPO infusion in CT2A mouse glioma. **a**, Immune phenotyping of ipsilateral cLN after IT PVSRIPO (extended data relating to Figure 1h). cLN weights, Tregs, $CD8^+$ $GzmB^+$ T cells, $CD11c^+$ MHC $II^+$ DCs and B cells in cLN 96h post mock IT vs. PVSRIPO IT treatment of CT2A glioma-bearing hCD155-tg mice. See Supplementary Figure 3b for gating strategy. **b**,**c**, Negative selection of MHC $II^+$ $CD11c^+$ DCs in cell suspensions from ipsilateral cLN 120h post IT PVSRIPO in CT2A mouse glioma (**b**). RT-qPCR results from total RNA isolated from whole cLN samples ('whole'), $CD11c^+$-bead isolated cells (vs. $CD11c^-$ fraction), and DC negative selection (vs. non-DCs) (**c**) (extended data relating to Figure 2a-c). **d**, Gating strategy for the identification of $CT^+$ DCs (extended data relating to Figure 2g).

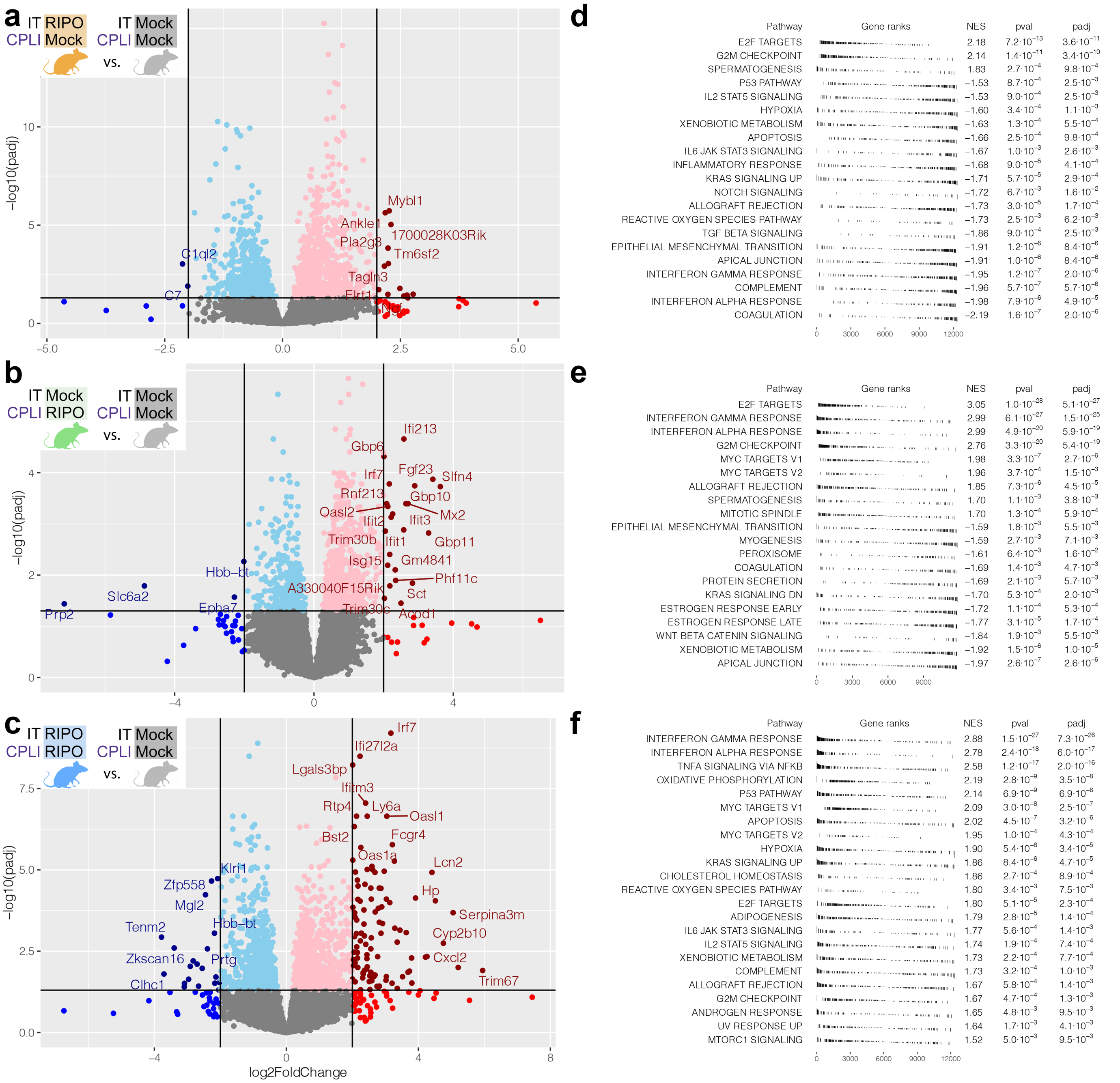


**g**

| | Description | Adjusted P Value | Gene Count | Fold Enrichment |
|---|---|---|---|---|
| *5* | protein methyltransferase activity | 2.48e−03 | 13 | 4.6 |
| *2* | histone modifying activity | 1.26e−04 | 22 | 3.6 |
| *3* | ribonucleoprotein complex binding | 1.43e−03 | 20 | 3.3 |
| *4* | catalytic activity, acting on DNA | 2.18e−03 | 22 | 3 |
| *1* | mRNA binding | 4.91e−06 | 36 | 2.9 |

**h**

| | Description | Adjusted P Value | Gene Count | Fold Enrichment |
|---|---|---|---|---|
| *6* | pattern recognition receptor activity | 1.01e−04 | 9 | 9.8 |
| *1* | double−stranded RNA binding | 1.49e−10 | 18 | 9.6 |
| *10* | histone H3 methyltransferase activity | 1.25e−03 | 8 | 8.8 |
| *9* | ubiquitin−like protein peptidase activity | 9.37e−04 | 13 | 5 |
| *3* | ribonucleoprotein complex binding | 4.56e−07 | 22 | 4.9 |
| *7* | nucleotidyltransferase activity | 2.42e−04 | 15 | 4.8 |
| *2* | catalytic activity, acting on RNA | 1.88e−10 | 36 | 4.2 |
| *5* | transcription coactivator activity | 1.48e−06 | 25 | 4.1 |
| *8* | modification−dependent protein binding | 3.33e−04 | 20 | 3.6 |
| *4* | ubiquitin−like protein ligase activity | 8.35e−07 | 30 | 3.6 |
| *11* | histone modifying activity | 7.42e−03 | 16 | 3.5 |
| *12* | microtubule binding | 8.89e−03 | 19 | 3 |
| *13* | DNA−binding transcription repressor activity, RNA pol II−specific | 2.32e−02 | 19 | 2.8 |
| *14* | GTP binding | 4.70e−02 | 21 | 2.5 |

**Supplementary Fig. 2.** Differences in gene expression and hallmark pathways between single-site (IT or CPLI) and dual-site (IT plus CPLI) treatment cohorts compared to mock treatment as shown. **a**-**c**, Volcano plots depict gene expression differentials when comparing treatment cohorts (extended data relating to Figure 3e). **d**-**f**, Gene set enrichment analysis (GSEA) hallmark pathways ranked by normalized enrichment scores (NES) (extended data relating to Figure 3f). **g**, **h**, Gene ontology overrepresentation of molecular function terms from significantly upregulated genes of IT (**g**) or CPLI (**h**) PVSRIPO treatment compared to the respective single-site (IT or CPLI) mock (extended data relating to Figure 3g).

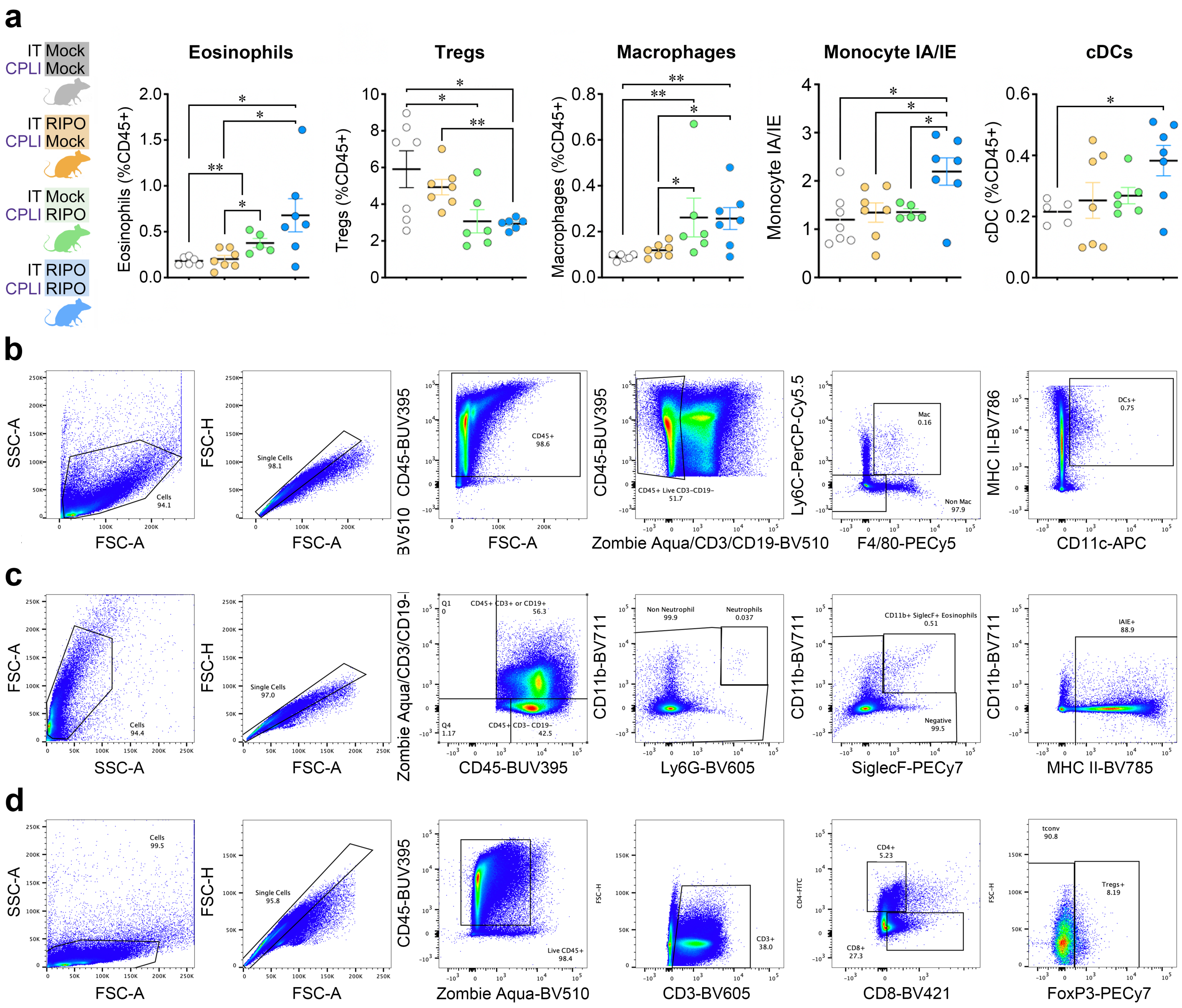


**Supplementary Fig. 3.** Dual-site (IT+CPLI) PVSRIPO in CT2A-bearing mice changes immune cell composition in cLN (extended data relating to Figure 3). **a**, Immune phenotyping in cLN from four PVSRIPO treatment cohorts as shown in Figure 3a, b. **b-d**, Gating strategy used for immune phenotyping of: macrophages and DCs in cLN (b); eosinophils and MHC II (IA/IE) expression on monocytes in cLN (c); T cells, including FoxP3$^{+}$ Tregs, in cLN (d).

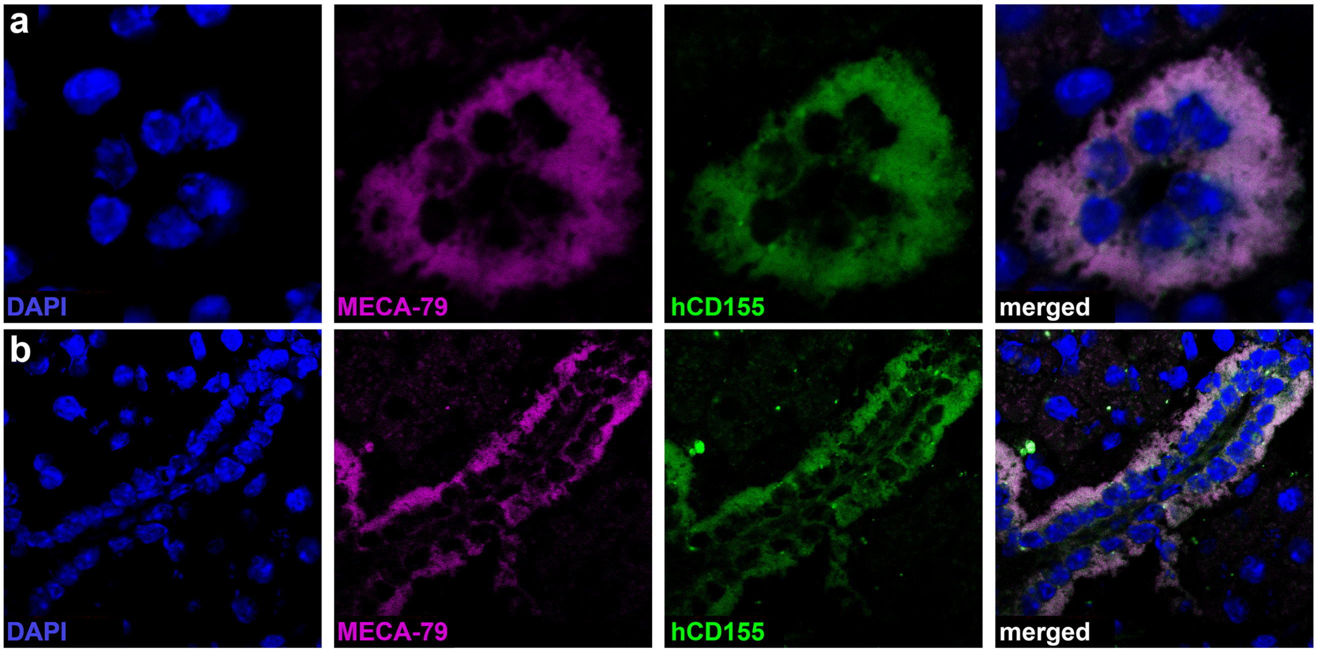


**Supplementary Fig. 4. a**,**b**, Extended data relating to Figure 4c-g. MECA-79$^{+}$ HEV, which harbor (-)strand vRNA in CT2A glioma-bearing mice after dual site (IT+CPLI) PVSRIPO, stain positive for the poliovirus receptor CD155.

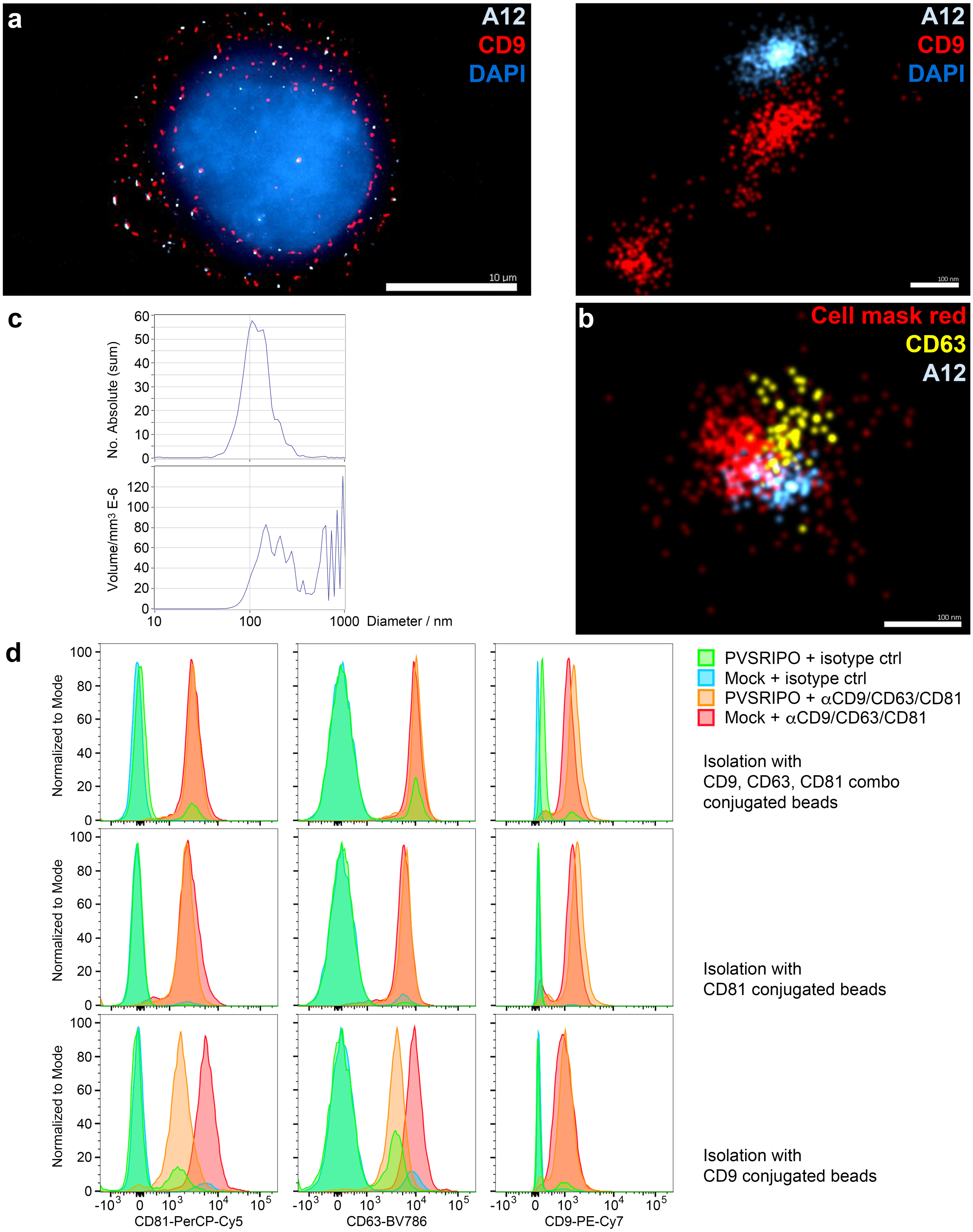
a
A12
CD9
DAPI
10 µm
100 nm
b
Cell mask red
CD63
A12
100 nm
c
No. Absolute (sum)
Volume/mm3 E-6
Diameter / nm
d
PVSRIPO + isotype ctrl
Mock + isotype ctrl
PVSRIPO + αCD9/CD63/CD81
Mock + αCD9/CD63/CD81
Isolation with
CD9, CD63, CD81 combo
conjugated beads
Isolation with
CD81 conjugated beads
Isolation with
CD9 conjugated beads
Normalized to Mode
CD81-PerCP-Cy5
CD63-BV786
CD9-PE-Cy7

**Supplementary Fig. 5.** PVSRIPO is shed in phosphatidyl-serine enriched vesicles from A375 melanoma cells (extended data relating to Figure 4g). **a**, Direct Stochastical Optical Reconstruction Microscopy (dSTORM) of an A375 cell infected with PVSRIPO (48hpi) stained for A12 (polio capsid antibody that only recognizes intact viral particles) and the tetraspanin CD9 (left panel). The high magnification detail depicts co-localization of A12 and CD9 stain (right panel). **b**, dSTORM depicts an extracellular vesicle (isolated from PVSRIPO-infected A375 cells) containing a PVSRIPO particle. Vesicles were isolated by differential centrifugation and $CD9^{+}$-bead isolation from A375 cell culture supernatants 48hpi with PVSRIPO. The sample was stained for cell mask red (marking plasma membrane), tetraspanin CD63, and A12. **c**, Nanoparticle Tracker results showing the distribution of diameters (top) and volumes (bottom) of vesicles isolated from PVSRIPO-infected A375 cells as described for **b**. **d**, Vesicles from mock- or PVSRIPO-infected A375 cells were isolated with CD81-, CD9- or a cocktail of CD81/CD9/CD63-conjugated beads, labeled with anti-CD81/9/63 or isotype controls, and tested for the expected vesicle markers/tetraspanins CD81, CD9 and CD63.

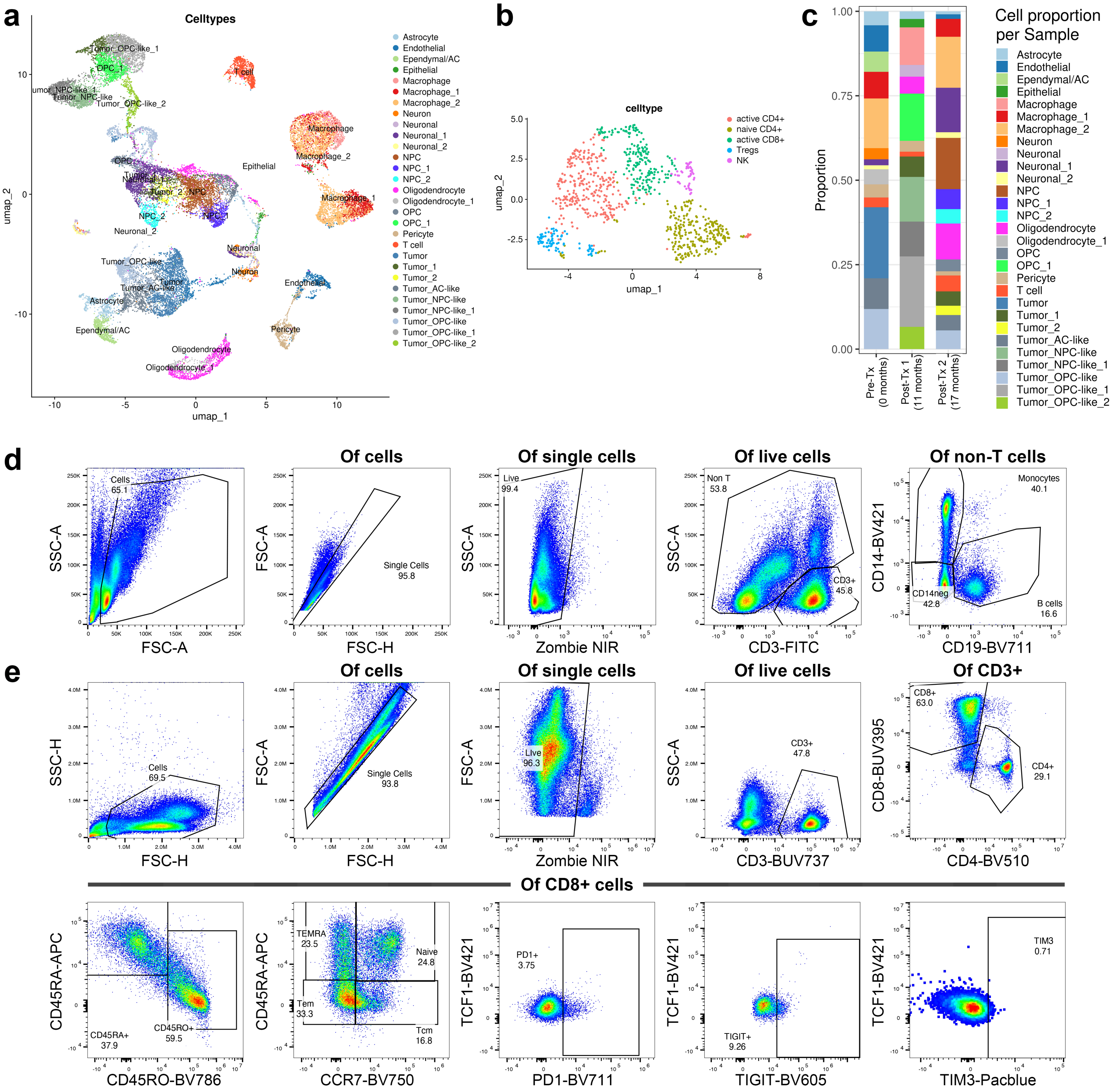


**Supplementary Fig. 6. a-c**, Extended data relating to Figure 5. Umap of snRNA seq of combined samples from three intervals (see Figure 5d); individual cell types are labeled (**a**). Umap of cells identified as T cells in **a**; individual subsets are labeled (see Supplementary Table 2 for extended data) (**b**). Proportion of cell types identified in **a** by time point (**c**). **d**, **e**, Gating strategy used (extended data relating to Figure 5e, f).
